# Monitoring of SARS-CoV-2 variant dynamics in wastewater by digital RT-PCR : from Alpha to Omicron BA.2 VOC

**DOI:** 10.1101/2022.04.04.22273320

**Authors:** Sebastien Wurtzer, Morgane Levert, Eloïse Dhenain, Heberte Accrombessi, Sandra Manco, Nathalie Fagour, Marion Goulet, Nicolas Boudaud, Lucie Gaillard, Isabelle Bertrand, Julie Challant, Sophie Masnada, Sam Azimi, Miguel Guillon-Ritz, Alban Robin, Jean-Marie Mouchel, OBEPINE SIG, Laurent Moulin

## Abstract

Throughout the COVID-19 pandemic, new variants have continuously emerged and spread in populations. Among these, variants of concern (VOC) have been the main culprits of successive epidemic waves, due to their transmissibility, pathogenicity or ability to escape the immune response. Quantification of the SARS-CoV-2 genomes in raw wastewater is a reliable approach well-described and widely deployed worldwide to monitor the spread of SARS-CoV-2 in human populations connected to sewage systems. Discrimination of VOCs in wastewater is also a major issue and can be achieved by genome sequencing or by detection of specific mutations suggesting the presence of VOCs. This study aimed to date the emergence of these VOCs (from Alpha to Omicron BA.2) by monitoring wastewater from the greater Paris area, France, but also to model the propagation dynamics of these VOCs and to characterize the replacement kinetics of the majority populations. These dynamics were compared to various individual-centered public health data, such as regional incidence and proportions of VOCs identified by sequencing of isolated patient strains. The viral dynamics in wastewater highlighted the impact of the vaccination strategy on the viral circulation in human populations but also suggested its potential effect on the selection of variants most likely to be propagated in immunized populations. Normalization of concentrations to capture population movements appeared statistically more reliable using variations in local drinking water consumption rather than using PMMoV concentrations because PMMoV fecal shedding was subject to variability and was not sufficiently relevant in this study. The dynamics of viral spread was observed earlier (about 13 days on the wave related to Omicron VOC) in raw wastewater than the regional incidence alerting to a possible risk of decorrelation between incidence and actual virus circulation probably resulting from a lower severity of infection in vaccinated populations.

## Introduction

The COVID-19 pandemic, caused by severe acute respiratory syndrome coronavirus 2 (SARS-CoV-2), is marked by the continuous emergence of numerous variants (World Health Organization, 2022). Among these, some variants show clear evidences of increased transmissibility, pathogenicity or better ability to escape human immune response induced by natural infection or vaccination (Cele et al., 2022, 2021; Planas et al., 2021; Wibmer et al., 2021). The identification of these variants, designated as variants of concern (VOC) by the WHO (World Health Organization, 2022), is of great importance with regard to vaccine strategy, use of certain therapeutic approaches based on neutralizing monoclonal antibodies, to allow a better understanding of the outbreak dynamics and, more generally, the definition of health policies decided by governments.

Over the course of the pandemic, different waves of infections have been reported, each of which was mainly associated with one or more VOC. Currently, five SARS-CoV-2 variants were designated as VOCs (Covariants.org, 2022; World Health Organization, 2022). The Alpha VOC (Pango lineage B.1.1.7; clade 20I) became the dominant SARS-CoV-2 variant worldwide in early 2021. At the same time, the Beta (lineage B.1.351; clade 20H) and Gamma (lineage P.1; clade 20J) variants were also identified around the world. From May 2021, the Delta variant (lineage B1.617.2, clade 21A) subsequently became dominant and was mainly responsible for the fourth wave observed in France during the last quarter of 2021. At the end of 2021, the Omicron variant (lineage B.1.1.529) was identified in multiple European countries and was designated as a VOC in November 2021 (European Centre for Disease Prevention and Control, 2022). Recently, 3 sub-lineages of Omicron VOC were described based on phylogenetic analysis and were designed BA.1 (clade 21K, the former that defined the Omicron lineage), BA.2 (clade 21L) and BA.3 very minor to date (Covariants.org, 2022). Omicron VOC has a panel of mutations associated with enhanced transmissibility and high potential for vaccine-induced immune escape (Cele et al., 2022). In contrast to previous variants, Omicron VOC is thought to have a lower overall severity in vaccinated populations (Ferré et al., 2022; Maisa et al., 2022). However, Omicron VOC has been reported to induce moderate infections in younger populations, with a notable increase in multi-systemic inflammatory syndrome cases according to French health authorities (Santé publique France, 2022a). Omicron BA.1 VOC has led to an unprecedented global resurgence of infection (7-day rolling incidence > 5,000 cases /100,000 inhabitants) during the delta wave in December 2021 in France (Santé publique France, 2022b). In this regard, early warning of Omicron emergence and surveillance of its sub-lineage dynamics must be implemented.

In order to meet this high demand for screening, the French testing strategy has changed significantly since January 2022. Thus, antigenic tests alone have become sufficient to make a diagnosis of COVID-19 infection, without confirmation by RT-PCR analysis. Although this approach has made it possible to streamline the workload of medical analysis laboratories in towns and hospitals, the possibilities of performing molecular analyses on these samples, screening by PCR for specific mutations of interest and sequencing, have been greatly affected, leading to a bias in patient recruitment and a potentially partial view of the overall viral circulation and the prevalence of each variant. Identification of the infection dynamics and involved VOCs is of great importance, but if the variants become less and less virulent, the clinical epidemiology may no longer correlate with true viral circulation in the populations.

Since the beginning of the pandemic, a complementary epidemiological approach based on fecal shedding of virus and its detection in wastewater has been proposed (Medema et al., 2020a). Although many methodological approaches have been described and a more standardized protocol is required (Ahmed et al., 2021; Bivins et al., 2021b, 2021a), the add-value of this approach has been widely demonstrated through the monitoring of the global viral burden of SARS-CoV-2. In this regard, the European Commission recommendation (EU) 2021/472 of 17 March 2021 called for the establishment of a systematic and harmonized surveillance of the SARS-CoV-2 genome and its variants in raw wastewaters in the European member states (Official Journal of the European Union, 2021).

The establishment of such a surveillance system is of significant interest in a context of low viral circulation to alert on potential viral emergence or re-emergence. The monitoring of variants, especially VOC, can also be performed in raw wastewater but with some limitations. Sequencing, the most frequently described method (Agrawal et al., 2022a; Crits-Christoph et al., 2021; Fontenele et al., 2021; Pérez-Cataluña et al., 2021; Rios et al., 2021), is complex to perform in wastewater due to the relatively low viral load compared to clinical samples and the potentially degradation and/or fragmentation of the viral RNA in such samples. In addition, the presence of multiple lineages in raw wastewater does not allow the proper assembly of reads to determine the complete sequences of circulating genomes and correct lineage assignment without a complex workflow. Even if the data quantity generated by sequencing is relevant, this methodology is also expensive and time-consuming. To overcome these limitations, targeted and specific RT-qPCR approaches have been proposed to detect certain mutations suggestive of variant presence or functional mutations whose evolution could reveal the spread of variants designated as VOC (Wurtzer et al., 2022). Different RT-PCR methods have been proposed based on either relative quantification of mutations to the total SARS-CoV-2 genomes (Erster et al., 2022; Wurtzer et al., 2022; Yaniv et al., 2021), either on wild-type and mutated allelic discrimination (Graber et al., 2021), quantification of mutations and total genomes by digital PCR (Boogaerts et al., 2022; Caduff et al., 2022; Heijnen et al., 2021; Lou et al., 2022). These approaches have been developed and applied to the emergence of Alpha and Delta VOC. To our knowledge, few data are available regarding the emergence and the dynamics of the Omicron BA.1 and BA.2 VOC (Agrawal et al., 2022b; Ahmed et al., 2022; Chassalevris et al., 2022; Kirby et al., 2022; Smyth et al., 2022).

In this study, the specific monitoring of VOC-suggestive mutations was performed by digital RT-PCR. The objectives were to date the emergence of the Omicron BA.1 and BA.2 sub-variants that have been recently detected in France, but also to model the propagation dynamics of these specific mutations in comparison with open statistical data from the national sequencing program (EMERGEN) of clinical samples (Santé publique France, 2022b). Population replacement kinetics of dominant variants (from Alpha to omicron BA.2) was determined. A comparison of the mutation proportions determined by digital RT-PCR and sequencing of wastewater samples was also performed. In addition, methods of data normalization, including human fecal biomarker, were tested on the data set.

## Material & methods

### Sample collection

Eight wastewater treatment plants (WWTP) were sampled twice a week in greater Paris area and 7 in other French regions. Sixteen sewers were weekly sampled in Paris, France (See map in Supplementary Figure 1). Twenty-four-hours composite samples (according to NF T 90-90-523-2) were taken by automated samplers. Sampling was based on flow rate, started at 7:00 AM and finish at J+1, 7:00 AM. Samples were taken by suction and collected in a refrigerated polyethylene tank at 5°C (+or-3°C). The final collected volume was between 8.7 and 14L. Then samples were carefully homogenized, distributed in a 100mL polyethylene bottle, transported to the laboratories at 4°C and processed in less than 24 hours after sampling.

**Figure 1.**
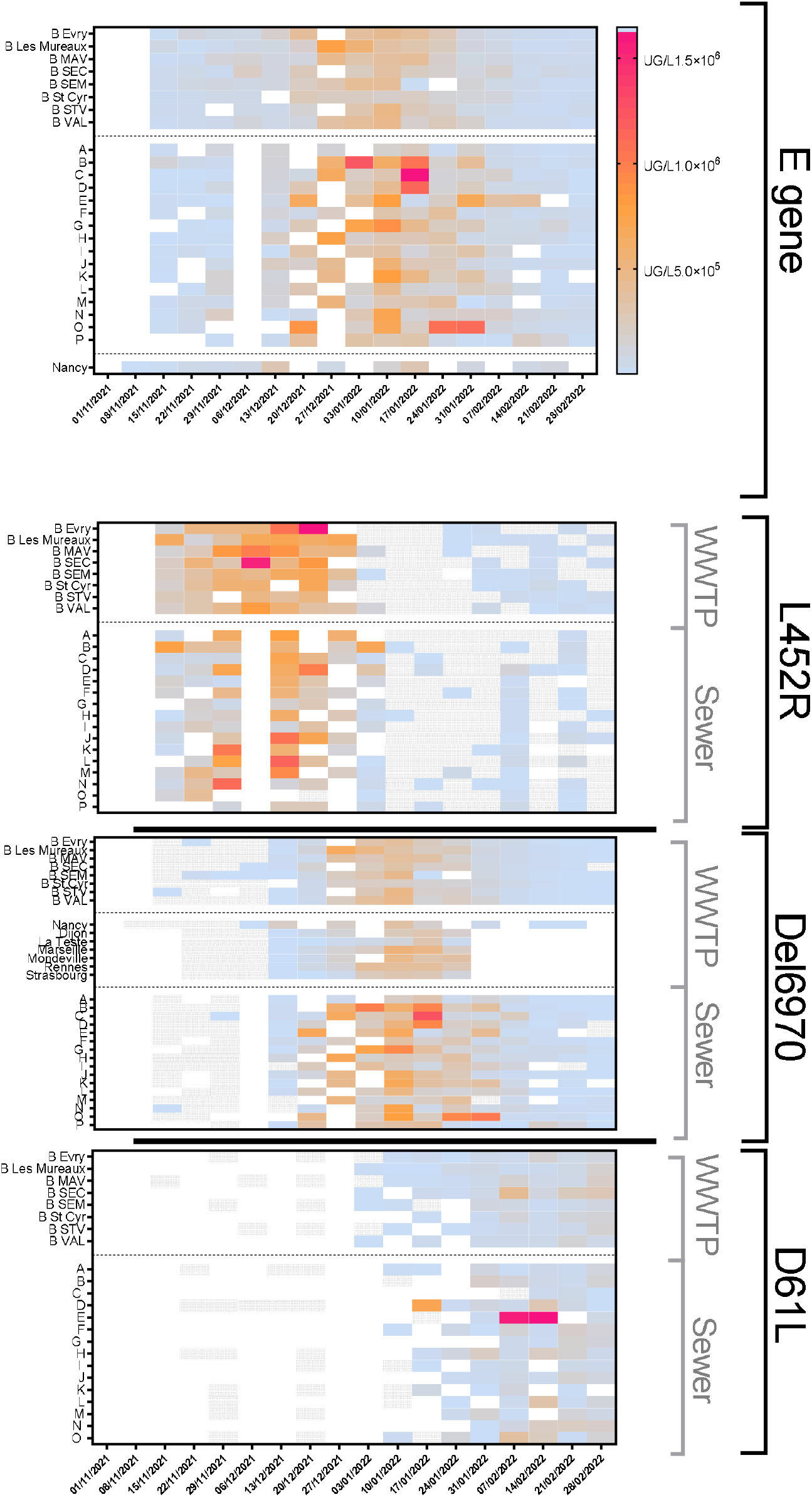
Early detection of VOC-evocative mutation in the greater Paris area and other French regions from mid-November, 2021 to mid-March, 2022. Wastewater treatment plants (WWTP) and sewers in Paris, France were sampled twice a week. Heat-maps showed the concentration of total SARS-CoV-2 genomes and those carrying specific mutations (L452R mutation for Delta VOC, del69-70 for Omicron BA.1 VOC, and D61L for Omicron BA.2 VOC). White box meant not analyzed, gray box meant not detected.

A total of 625 samples from the greater Paris area were analyzed and 72 from other French regions.

### Concentration methods

All samples from Paris greater area, including WWTP and sewer samples, were processed as previously described (Wurtzer et al., 2020). Briefly, samples were homogenized, then 11 ml were centrifugated at 200 000 x g for 1 hour at +4°C using a XPN80 Coulter Beckman ultracentrifuge using a swing rotor (SW41Ti). Pellets were resuspended in 200 μL of PBS 1X buffer and pretreated for dissociating viruses and organic matter that was then removed from supernatant for improving RNA extraction efficiency, according to the manufacturer’s recommendations. Supernatant was then lysed, and total nucleic acids were purified using PowerFecal Pro kit (QIAGEN) on a QIAsymphony automated extractor (QIAGEN) and eluted in 50 µL of elution buffer according to manufacturer’s protocol.

The other samples from WWTP were processed according to an alternative protocol (Bertrand et al., 2021). Briefly, 5mL of raw wastewater were used for nucleic acid extraction using 10 mL of NucliSENS® lysis buffer. A purification step using phenol/chloroform/isoamyl alcohol 25:24:1 could be performed depending on the laboratories. The nucleic acid extraction was continued using 70 μL of magnetic silica beads and the NucliSENS® platforms (easyMAG™ or miniMAG™) (BioMérieux). The nucleic acids were eluted in 50 μL of elution buffer.

All nucleic acids were finally purified using OneStep PCR inhibitor removal kit (Zymoresearch) according the manufacturer’s instructions and then stored at -80°C until molecular assays.

The recovery rate of methods was estimated using bovine coronavirus spiked and the repeatability of the measurement was also evaluated on endogenous PMMoV as previously described (Wurtzer et al., 2022).

### Molecular quantification by digital RT-PCR

A panel of oligonucleotides was designed using AlleleID software vers. 7 (Premier Biosoft) in order to detect and quantify mutations reported in table 1. Briefly, Alpha and Omicron BA.1 VOC have been reported for sharing a deletion of amino acids H69 and V70 in the spike protein (del 69-70), but Omicron BA.1 VOC can be distinguished from Alpha by an additional synonymous substitution (C/T) at the position 21762 (codon 67 of the spike protein). Delta VOC has been reported carry the L452R mutation in the spike protein. Bioinformatics analyses revealed that Omicron BA.2 variant carried the D61L mutation in the ORF6. In addition, PMMoV genome was also detected using previously described oligonucleotides (table 1).

**Table 1.**
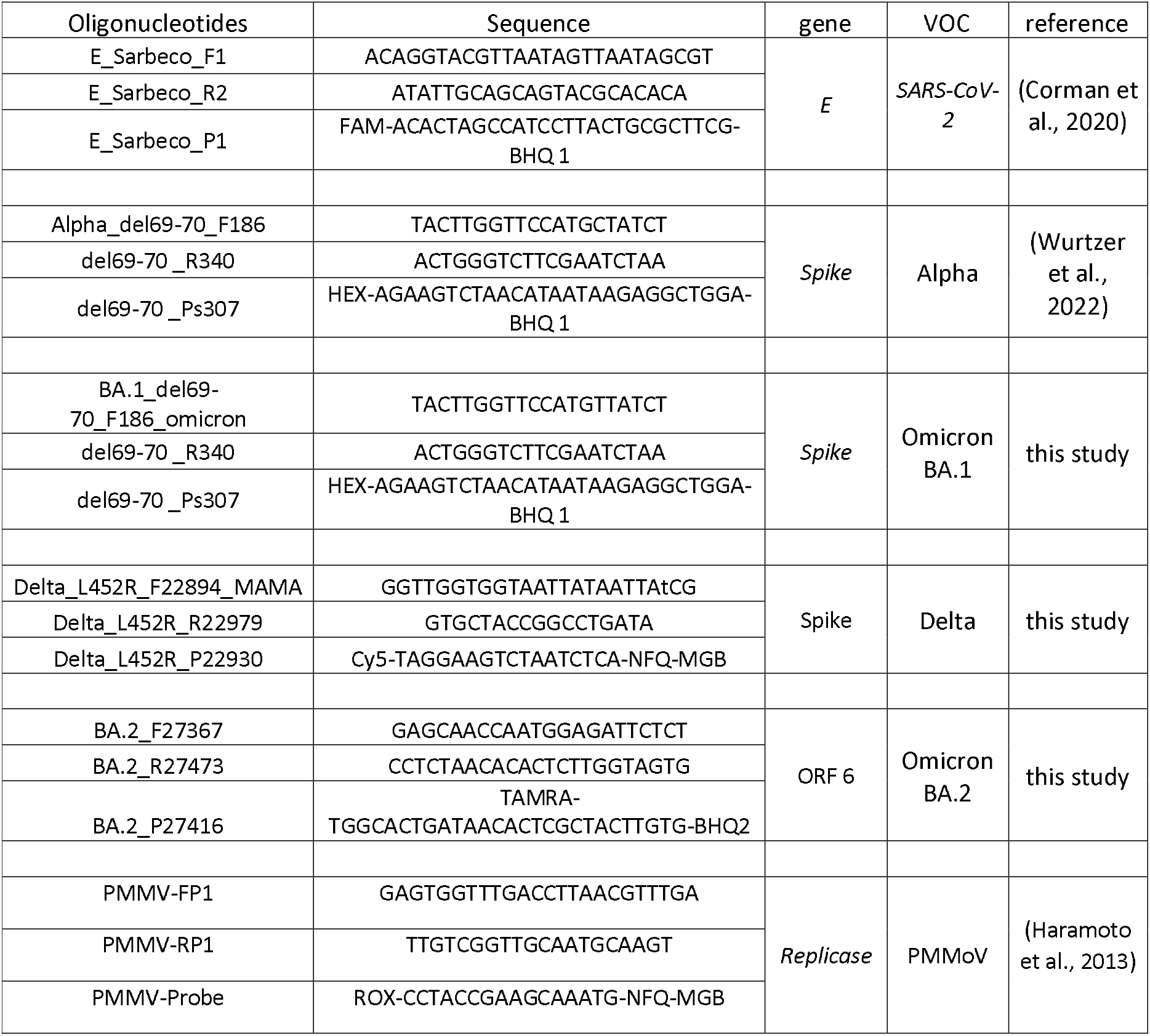
Oligonucleotides used for detecting SARS-CoV-2 and VOCs

Briefly, total SARS-CoV-2 genomes and mutations were quantified by digital RT-PCR. All samples from greater Paris area were processed using a QIAcuity instrument (QIAGEN). Multiplex reactions were performed using QIAcuity One-Step Viral RT-PCR mastermix (QIAGEN) with 800 nM of each primer and 250 nM of each probe in 26k-Nanoplates according to the manufacturer’s recommendations (table 1). In order to improve the detection specificity of single nucleotide polymorphism, the thermal profile included a touchdown step. Briefly, reverse transcription was done at 50°C for 10 min, followed by activation step at 95°C for 2 min. Then touchdown was composed by 5 cycles including a 5 sec-denaturation step at 95°C followed by a hybridization/elongation step from 63°C with a decrease of 1°C/cycle for 40 sec. Amplification was done by 45 cycles of incubation at 95°C for 5 sec and 58°C for 40 sec. The use of these nanoplates provided a minimum of 24k valid partitions for each sample and an estimated detection limit of 1800 copies/L.

Other samples were only processed for the quantifying del69-70 mutation among total genomes using QX200 instrument (BioRad). The ddRT-PCR assays were performed in a 20 μL-reaction mixture containing 5 μL of extracted nucleic acids and 15 μL of One-Step RT-ddPCR™ Kit for Probes (Bio-Rad). The reaction mix contained 909 nM of each primer and 227 nM of probe. The samples were placed in the droplet generator using 70 μL of generator oil. The resulting picolitre droplet emulsions (40 μL) were transferred to a T100 Thermal Cycler or CFX96 (Bio-Rad). The reverse transcription was performed with 60 min hold at 50°C. The cDNA amplification was performed with 10 min hold at 95°C, 40 cycles of 95°C for 30 sec then 58°C for 60 sec with a ramp rate of 2°C per sec, followed by 10 min hold at 98°C and 30 min hold at 4°C, and finally the maintain of the samples at 12°C until data analysis. After amplification, the plate was transferred to QX200 Droplet Reader (Bio-Rad) using QX Manager Edition™ Software (Bio-Rad) to measure the number of positive and negative droplets based on fluorescence amplitude. The detection limit was estimated to 2.000 copies/L.

Negative controls were included in each experiment to ensure no contamination and set up the thresholds for considering a partition as positive. Detection specificity of variant was assessed in each reaction using positive RNA controls kindly provided by Pr. Charlotte Charpentier (University Hospital Center Bichat-C. Bernard, Paris, France) and Pr. Evelyne Schvoerer (CHRU Nancy, France).

### Sequencing of wastewater samples

Sequencing was performed based on targeted whole genome library preparation of SARS-CoV-2 using the QIAseq DIRECT SARS-CoV-2 kit (QIAGEN). Five microliters of extracted RNA from each sample were used to make this library. Amplicons were quantified using an ultra-sensitive fluorescent nucleic acid stain for quantitating double-stranded DNA (Quant-iT™ PicoGreen® dsDNA reagent, Thermo). Samples normalized at 4.0 nM were pooled into a single library and diluted to 10 pM concentration according to the manufacturer recommendations. Sequencing was performed with the MiSeq V2 chemistry with 2 × 150 cycles according to the manufacturer’s protocol. Data analysis was performed using the Galaxy tools. Briefly, no quality filtering was applied, all reads were considered. Reads were mapped with BWA-MEM (version 0.7.17.1) against reference genome (NC_45512). Duplicate were cleaned from the BAM dataset using Mark Duplicates (version 2.18.2.2). Variant calling was performed using Lofreq (version 2.1.5) and annotation of SARS-CoV-2 variants using SnpEff eff version 4.5 covid19).

### Mathematical and statistical methods

Dynamics of mutations (concentration or mutation frequency) were modeled based on daily average concentration (when multiple locations were sampled on the same day) using a LOWESS smoothing method on 10 adjacent points (GraphPad Prism v9.0), allowing to limit outlier effects. Correlations between different datasets were estimated using Spearman test (GraphPad Prism v9.0). First derivative of dynamics curves was calculated using dedicated function of GraphPad Prism software. Normalized data were calculated by dividing raw mutation concentrations by PMMoV concentrations or by daily drinking water consumption. To homogenize the scales of the graphics, the ratios were multiplied by 10.000.000 or 400.000 respectively.

## Results

### 1. Dating of the emergence of Omicron variants in France

The del 69-70 mutation of the VOC Omicron BA.1 spike protein and the L452R mutation carried by the Delta VOC were quantified by digital RT-PCR on wastewater samples as early as the first week of November 2021. Quantification of the D61L mutation in the ORF6, suggestive of the Omicron BA.2 variant, was implemented on the samples as early as mid-November 2021. The heat maps presented in Figure 1 highlighted a very early detection of del69-70 mutation in samples from the Greater Paris area as early as November 15, 2021 in a context of predominant Delta variant illustrated by the screening of the L452R mutation. A later detection of del 69-70 was observed in the other major French cities. Similarly, the D61L mutation was detected in samples from the greater Paris area as early as January 3, 2022 heralding the emergence and rapid spread of the Omicron BA.2 variant. The density of samples taken in the greater Paris area, both in wastewater treatment plants and sewage collectors of the city of Paris, allowed to observe at first punctual detections of these mutations, then the generalization of the positivity of the sampling sites was a precursor of the large diffusion of the BA.1 and BA.2 lineages of the Omicron VOC in the population and to the acceleration of their propagation.

### 2. Dynamics of mutation in wastewater and of variant based on patient sequencing

Two periods were considered and 625 samples were processed. The first period concerned the first half of 2021 during which the del69-70 and L452R mutations, carried by Alpha VOC and Delta VOC respectively, were quantified. The proportions of each mutation were estimated in comparison to the total SARS-CoV-2 genome concentrations based on gene E quantification. This period was marked by a strong predominance of del69-70 mutation suggesting Alpha VOC before decreasing from April 2021. It should be noted that the L452R mutation was detected as early as January 2021 and could be found in nearly 15% of the circulating genomes in the wastewater. Its increasing frequency from mid-April 2021 was accompanied by a decrease in the proportion of del69-70. In wastewater, the L452R mutation became the majority mutation by May 27, 2021. The increase in the proportion of the L452R mutation was strikingly associated with the regional dynamics of vaccination (first dose) (figure 2, panel A) (Santé publique France, 2022b). Nevertheless, the concentration of SARS-CoV-2 genomes in raw wastewater associated with this Delta VOC epidemic wave was greatly reduced compared to the Alpha VOC wave (figure 2, panel B). These mutation dynamics were compared to local open data from the national sequencing program (EMERGEN) as well (figure 2, panel A) (Santé publique France, 2022b). This program was only implemented in France in mid-February 2021 explaining the shape of the curve. The Delta VOC was reported for the first time through sequencing from patient samples on May 10, 2021. The Delta variant became the majority of clinical samples on June 21, 2021 ((Santé publique France, 2022b).

**Figure 2.**
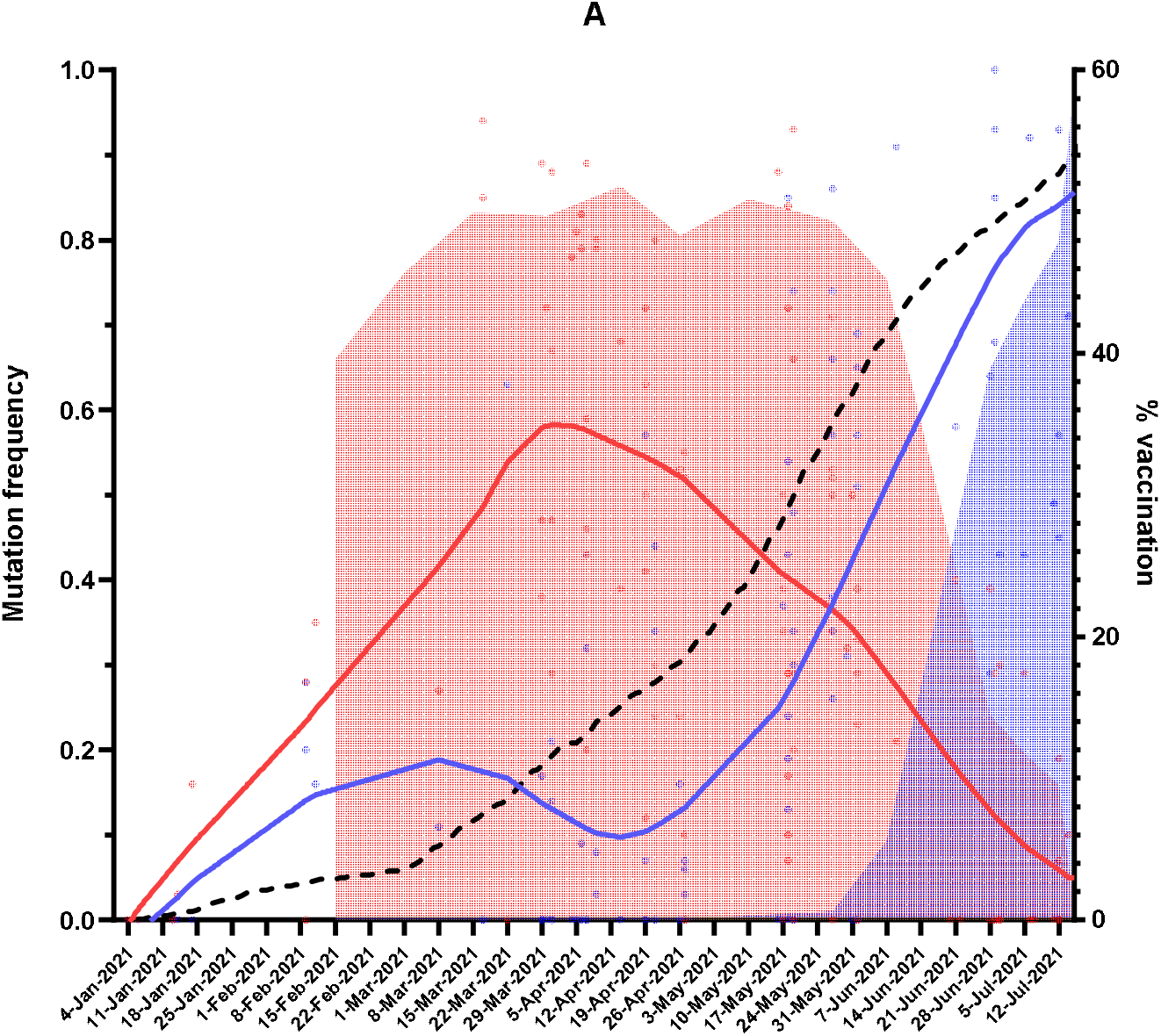
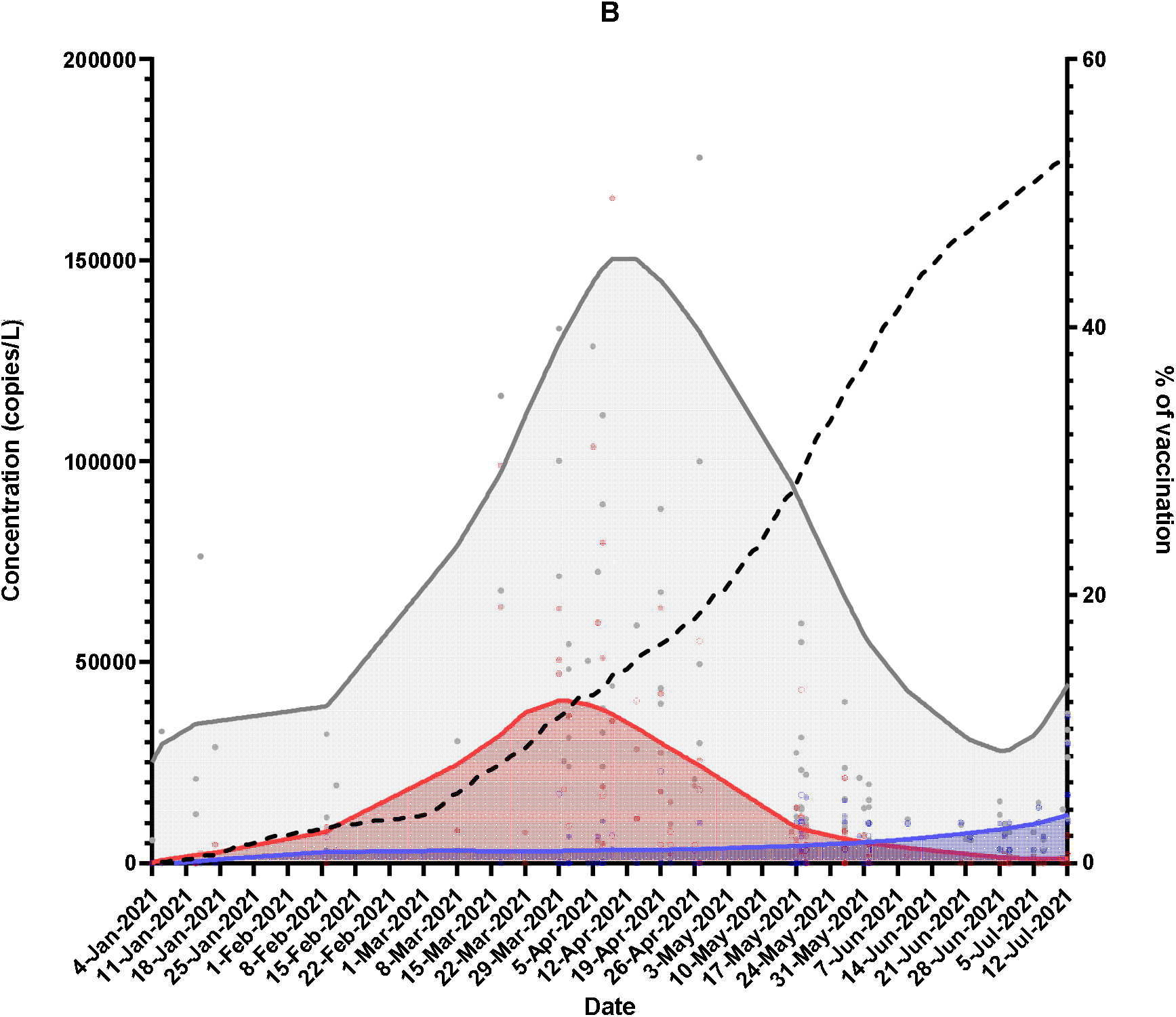
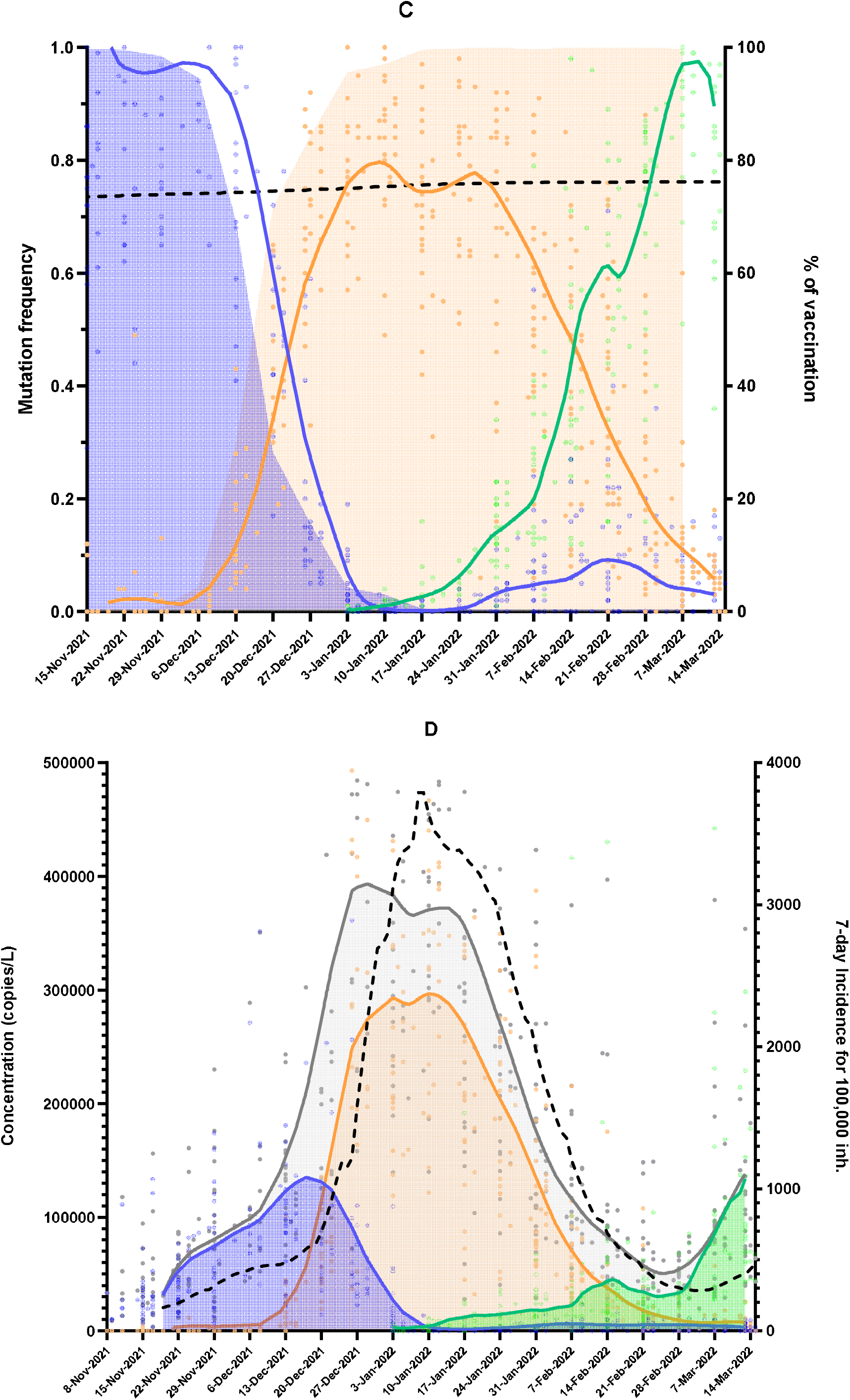
Dynamics of specific mutations in wastewater in greater Paris area from January 2021 to March 2022. Panel A: Frequency evolution of the del69-70 (red curve) and L452R (blue curve) mutations in wastewater from January 2021 to July 2021. The red area indicated the evolution of Alpha VOC by sequencing in clinical samples, the blue area for the evolution of Delta VOC. The dotted curve indicated the evolution of vaccination (percentage of people receving the first dose). Panel B: Concentration evolution of the del69-70 (red curve) and L452R (blue curve) mutations, and of the total SARS-CoV-2 genomes (grey curve) in wastewater from January 2021 to July 2021. The dotted curve indicated the evolution of vaccination (percentage of people receving the first dose). Panel C: Frequency evolution of the L452R (blue curve), del69-70 (orange curve) and D61L (green curve) mutations in wastewater from November 2021 to March 2022. The blue area indicated the evolution of Delta VOC by sequencing in clinical samples, the orange area for the evolution of Omicron VOC (distinction of BA.1 and BA.2 lineages was not available). The dotted curve indicated the evolution of the vaccination (percentage of people receving the first dose). Panel D: Concentration evolution of the L452R (blue curve), del69-70 (orange curve) and D61L (green curve) mutations, and of the total SARS-CoV-2 genomes (grey curve) in wastewater from November 2021 to March 2022. The dotted curve indicated the evolution of the regional incidence.

The second period began in November 2021, during which time Delta VOC represented 100% of identified viral sequences isolated from patients sequencing (Santé publique France, 2022b) and the L452R mutation was present in about 100% of circulating sequences in wastewater. The decrease of the L452R mutation in wastewater, concomitant with the decrease of the Delta VOC in clinical samples, started at the end of November 2021 and became a very small minority (<10% of sequences) by mid-January 2022. However, this L452R mutation was still detectable in wastewater by March 15, 2022 suggesting the circulation of variants carrying this mutation (Delta VOC or other non-VOC variants). The del69-70 mutation was once more detected in a wastewater sample from 15 November 2021 in the greater Paris area. Generalization and increase of this mutation were initiated at the end of November 2021 to become the majority on December 25, 2021. The growth dynamics of del69-70 in wastewater were very similar to that of the Omicron VOC estimated in patient by sequencing (Santé publique France, 2022b). In contrast, the del69-70 mutation began to decline by mid-January 2022 in parallel with the increase of the D61L (ORF6) mutation, detected in samples as early as January 3, 2022. Genomes carrying the D61L mutation became predominant by mid-February 2022 to present almost all circulating genomes by March 15, 2022 in the greater Paris wastewater. Sequencing results from patients did not report a distinction between Omicron BA .1 and BA.2 sublineages to date (Figure 2, panel C) (Santé publique France, 2022b). The association of the del69-70 mutation with the Omicron BA.1 VOC was based on a polymorphism in the forward primer and the absence of residual detection of this specific mutation in the summer of 2021 (data not shown). During this second period, the concentration of SARS-CoV-2 genomes was also compared to the regional incidence. The dynamics of the viral genome increase in wastewater that preceded the dynamics of incidence was of about 13 days (Figure 2, panel D). The concentration of viral genomes in wastewater resulting from the Omicron BA.1 VOC wave was much higher than those related to Delta VOC. To confirm the relevance of the digital RT-PCR method, the same samples were analyzed by RT-qPCR based on gene E quantification using the previously described method (Wurtzer et al., 2022). Trend curves were compared and showed close relationship (supplementary data 2).

### 3. Evolutionary trend: interest of raw concentration normalization

Viral concentrations in wastewater can be influenced by significant population changes since raw wastewater not solely comes from population (sewer cleaning, rain, water infiltration, industries…). To assess the impact of human population movements on the evolutionary trends of the different VOC mutations, comparison of raw mutation concentrations and normalized concentrations was performed using two indicators. The first one was related to an human biomarker, the Pepper Mild Mottle Virus genome (PMMoV), in wastewater. The second one was related to the consumption of drinking water by the local population living in Paris, France. PMMoV genome concentration and drinking water consumption were plotted in figure 3, panels A and B. Mean PMMoV genome concentration ranged between 10^6^ and 10^8^ copies/L with a mediane value of about 10^7^ copies/L, suggesting large fluctuations. On the contrary, the volume of drinking water consumed were more stable. An evolution of both parameters showed that a decrease in drinking water consumption was observed during holidays (shaded period on the graph) suggesting a decrease in the population connected to the watershed. Such variations were not observed with the PMMoV genome concentration.

**Figure 3.**
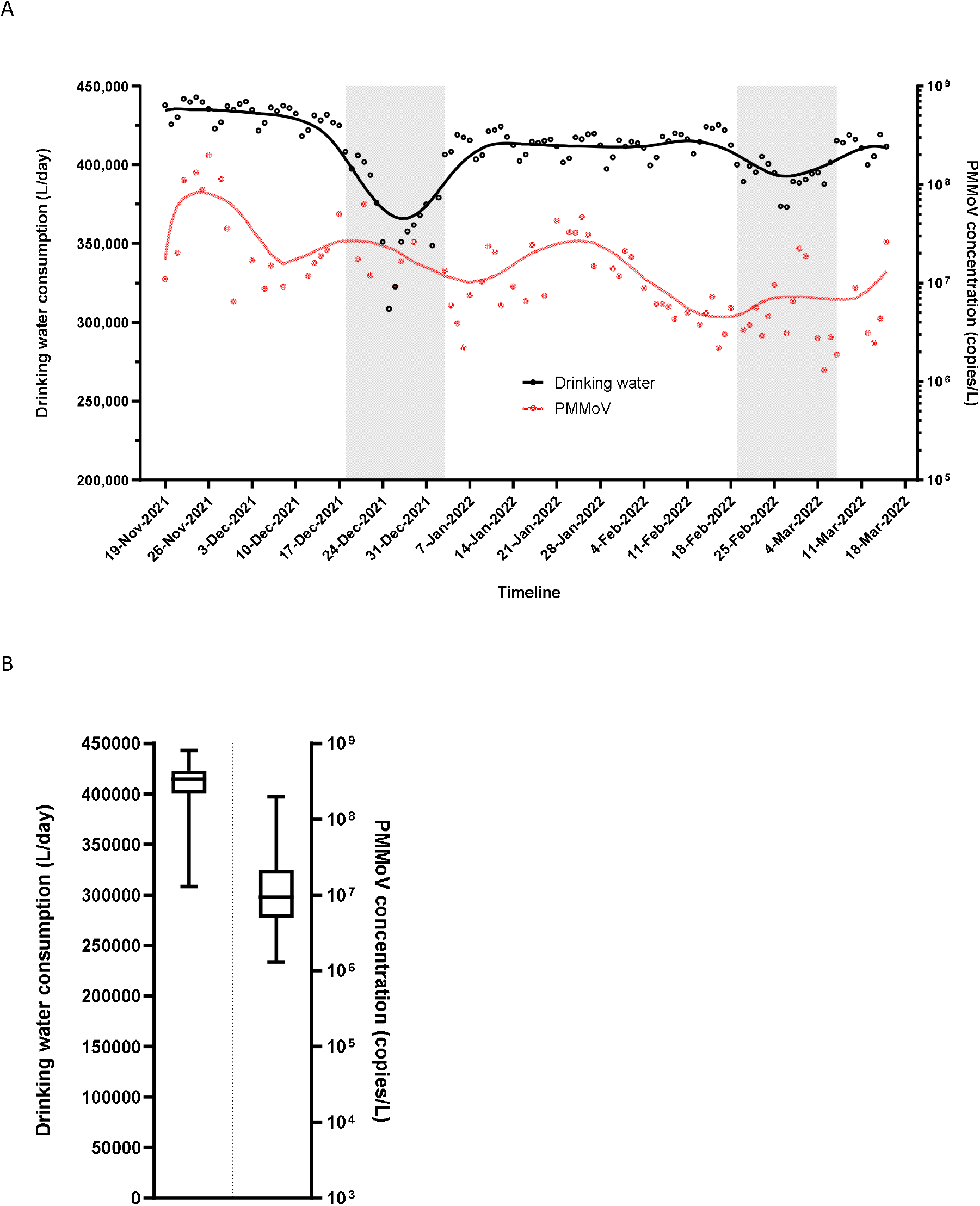
Evolution of human population indicators from November 2021 to March 2022. Panel A: Drinking water consumption in Paris, France (black curve) and PMMoV concentration in wastewater (red curve) were plotted, the grey areas indicated the vacation calendar. Panel B: Dispersion of drinking water consumption in Paris, France and PMMoV concentration in wastewater. Box and whiskers (minimum and maximum) were plotted.

Both datasets underwent the same LOWESS smoothing setup over the same period. The evolutionary trends of each mutation were very comparable, but some trend breaks can be observed (figure 4, panel D). Since the values were relatively stable (except during holydays), normalization by drinking water consumption only significantly altered the dynamics, mostly over the period of population movement (figure 4, panels A and B). Normalization with PMMoV noticeably altered viral dynamics in wastewater. The precocity of the viral dynamics was also reduced. This approach highlighted an epidemic rebound during February 2022 before an increase in March 2022 related to Omicron BA.2 (figure 4, panels A and C).

**Figure 4.**
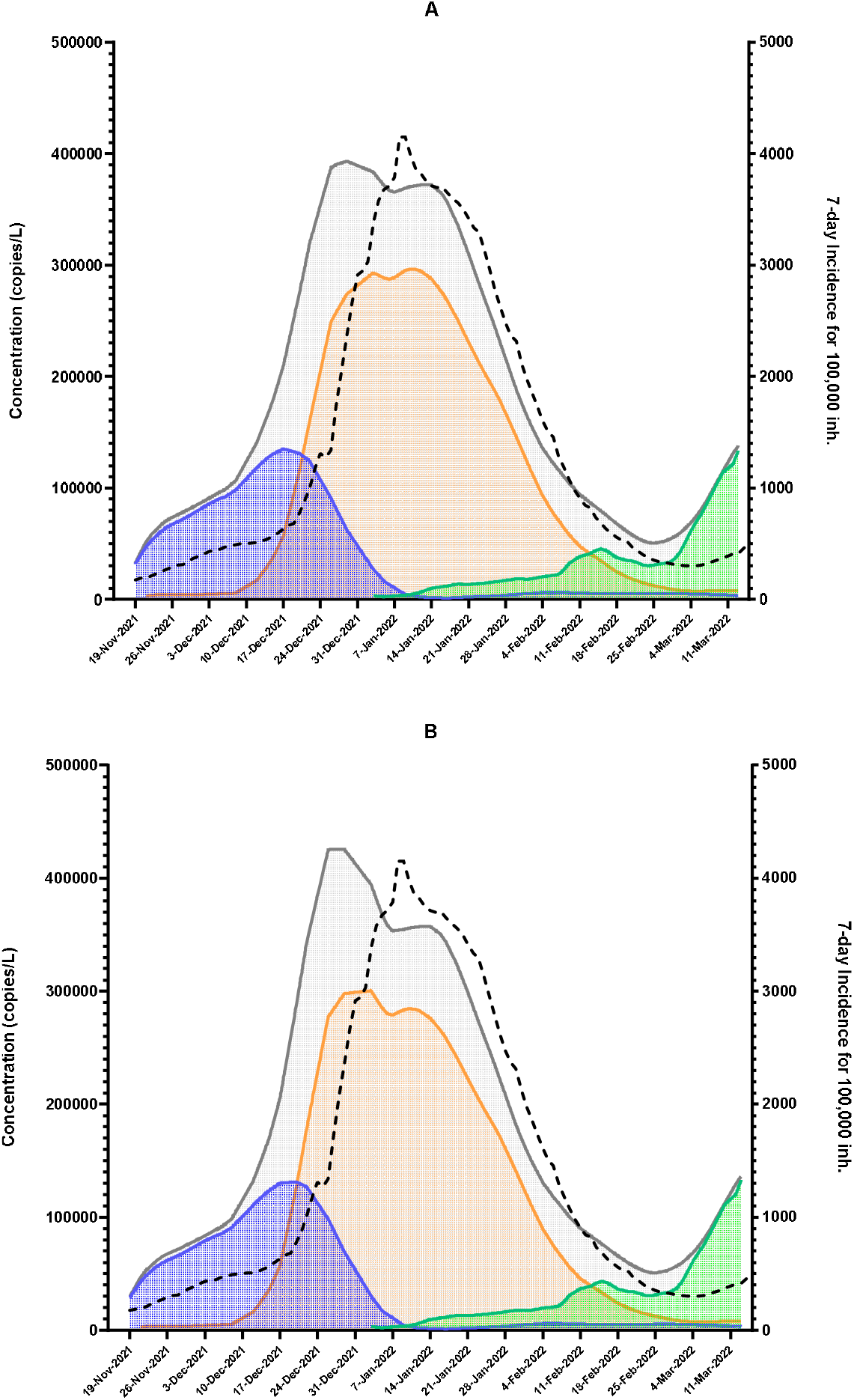
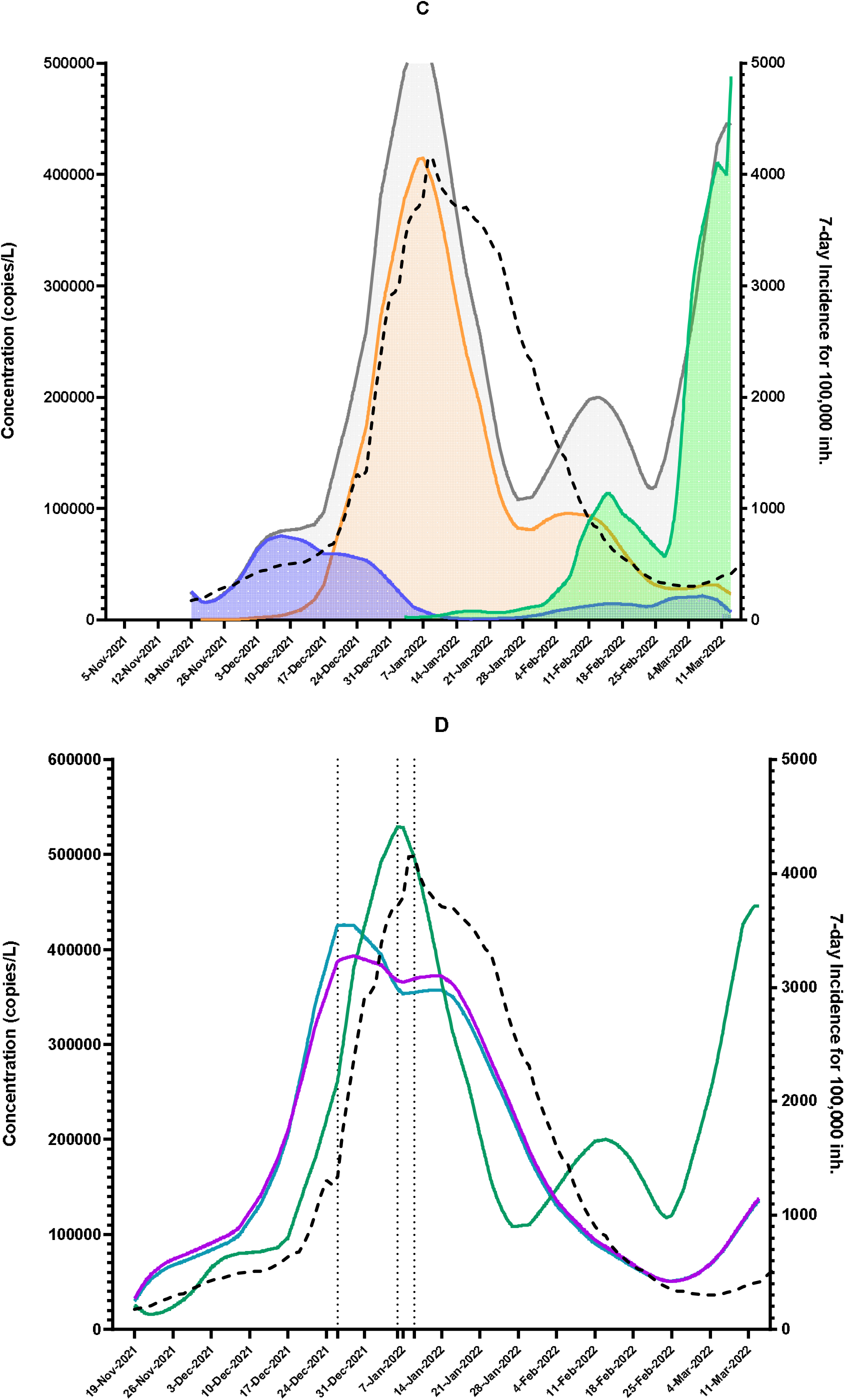
Evolutionary trends of the L452R (blue curve), del69-70 (orange curve) and D61L (green curve) mutation concentrations, and the total SARS-CoV-2 genome concentration (grey curve) in wastewater in greater Paris area from November 2021 to March 2022. The dotted curve indicated the evolution of the regional incidence. Panel A: trends based on raw concentrations. Panel B: trends based on concentrations normalized using drinking water consumption. Panel C: trends based on concentrations normalized using PMMoV concentrations measeared in wastewater. Panel D: Comparison of these 3 models and the incidence curve

The viral dynamics in raw wastewater having the best correlation with incidence data was determined by a Spearman correlation test. Taking into account the precocity of the raw and normalized curves using the drinking water consumption, these two modeling showed a very good correlation with the incidence curve (r = 0.956; p < 0.0001 and r =0.968; p < 0.0001) respectively, while the correlation of the normalized curve using the PMMoV genome concentration seemed to be less relevant (r = 0.761 ; p < 0.0001).

Normalization methods did not change the mutation frequencies and by the way the dynamics, but slight modifications of mutation concentrations were observed (figure 4, panels A, B and C).

### 4. Dynamics of variant proportions in wastewater: from Alpha to Omicron BA.2

The evolutionary kinetics of each mutation suggestive of VOC were studied over the 2 time periods. As PMMoV concentrations were not available for the first period and normalization by drinking water consumption did not significantly modify the modeling of viral dynamics, first derivatives of the raw mutation concentration trend curves were calculated to determine variant replacement rates based on the mutations studied. The values are presented in Table 2. The values corresponded to maximum rates of change in the proportion of mutation per day relative to the next majority population. Thus, the proportion of the del69-70 mutation of Alpha VOC increased by a maximum of 2.8% per day, compared with 5.2% for the L452R mutation of Delta VOC. The del69-70 mutation of Omicron BA.1 VOC increased by 6.0% per day in wastewater, while the D61L mutation replaced it more rapidly (maximum 17.2%/day) as soon as the wave started (March 2022). The rate of decay of the del69-70 mutation of Omicron BA .1 VOC was similar to those of L452R mutation.

**Table 2.**
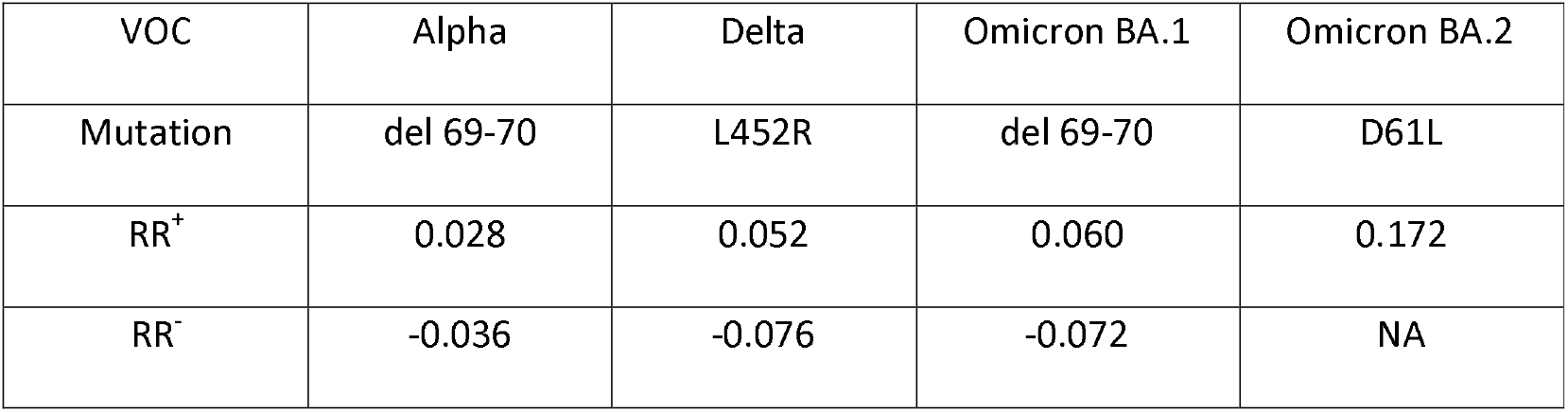
Maximal replacement rate (RR) of variant in wastewater. RR+ indicated the replacement rate during the increase of mutation frequency, whereas RR-provided information on the decay rate of the frequency. Values were indicated in increase or decrease of frequencies per day.

The sum of the concentrations of the mutations of interest carried by the different genomes was compared with the total concentration of circulating genomes over the two periods studied. During the first period, the correlation between both datasets was estimated using a Spearman test (r = 0.789 ; p < 0.0001) and a shift with the SARS-CoV-2 genome curve was observed suggesting the circulation of other variants than Alpha and Delta circulating in the wastewater (figure 5, panel A). In the second period, the sum of variants carrying the L452R, del69-70 and D61L mutations appeared to be more consistent with the total concentration of circulating genomes, independently of the viral concentration in wastewater (r = 0.966 ; p < 0.0001) suggesting less « other variants » circulating in wastewater (figure 5, panel B).

**Figure 5.**
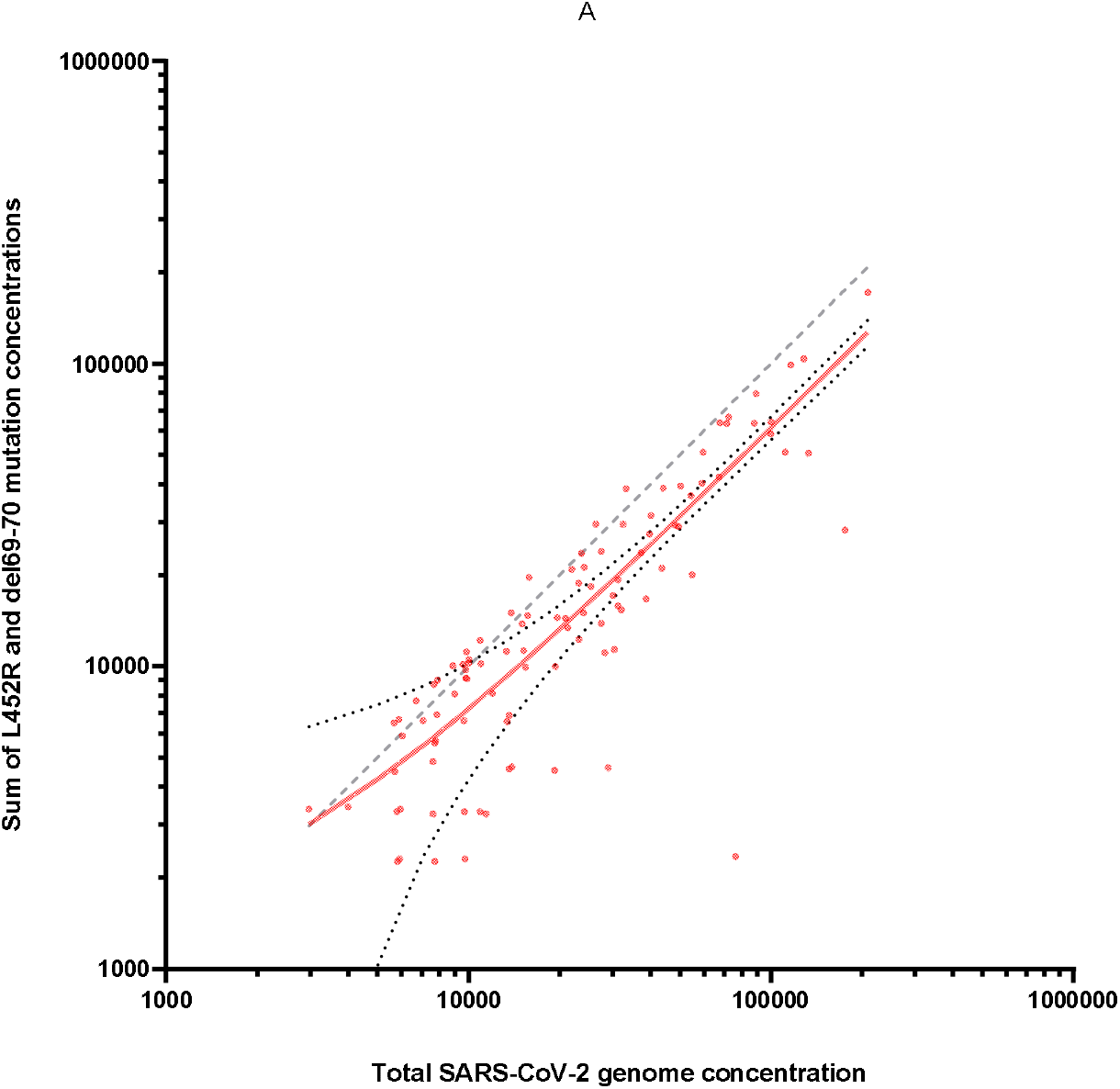
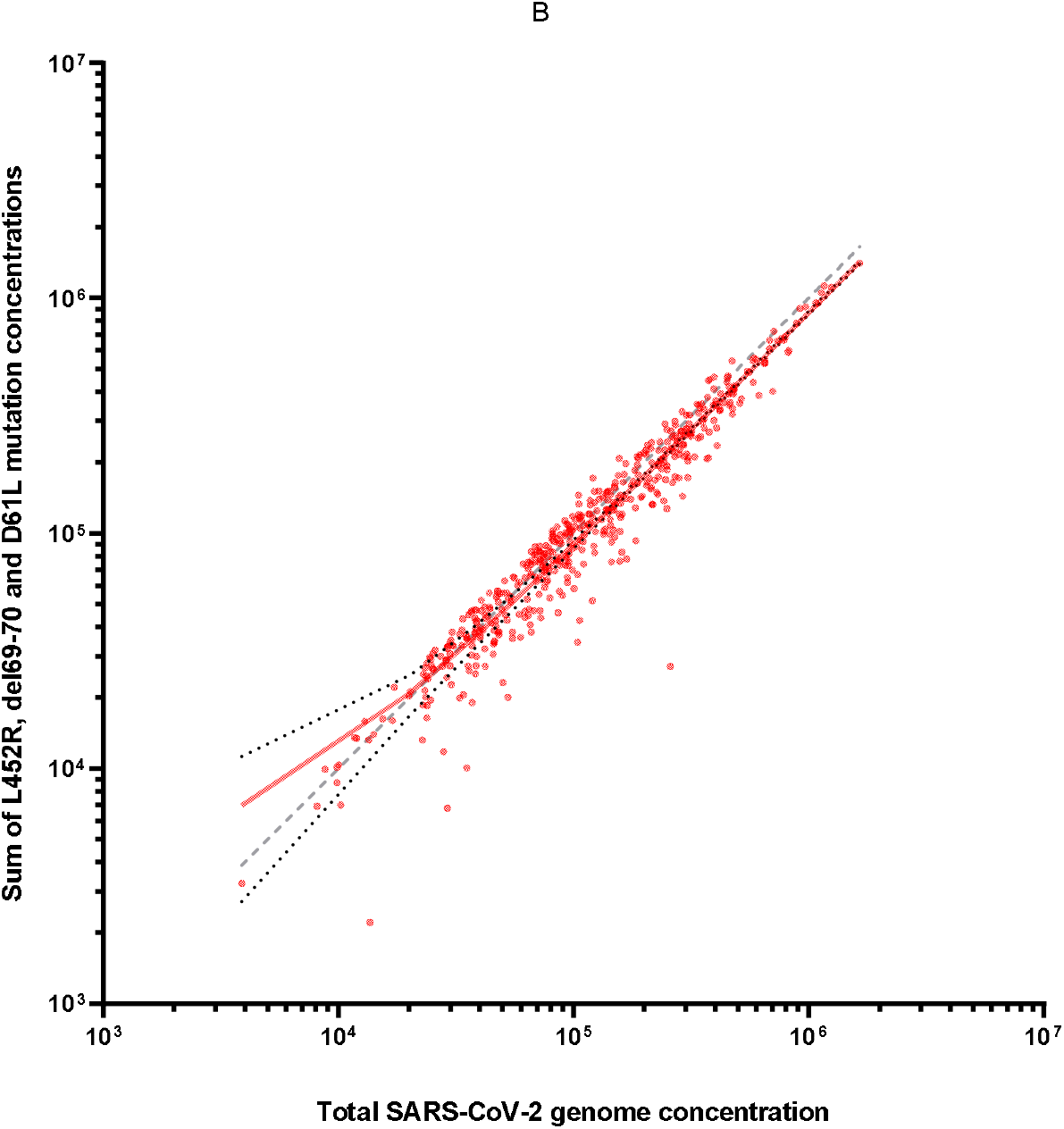
Correlation between the total SARS-CoV-2 genome concentration and the sum of VOC-suggestive mutations in wastewater in the greater Paris area. Panel A: Total SARS-CoV-2 genome concentrations were compared to the concentration sum of del69-70 and L452R mutations measured from January 2021 to July 2021. Grey dotted line indicated the theorical correlation whereas the red line was the linear regression and the 95% confidence bands of the best-fit line. Panel B: Total SARS-CoV-2 genome concentrations were compared to the concentration sum of del69-70, L452R and D61L mutations measured from November 2021 to March 2022. Grey dotted line indicated the theorical correlation whereas the red line was the linear regression and the 95% confidence bands of the best-fit line.

### 5. Comparison of mutation frequencies by digital RT-PCR and sequencing

The frequencies of del69-70, L452R, and D61L mutations were estimated by sequencing on 23 wastewater samples collected in the greater Paris area between December 15, 2021 and February 28, 2022. These mutation frequencies were compared to those estimated on the same samples by digital RT-PCR (Figure 6, panel A). The SARS-CoV-2 concentrations ranged from 3.3E^+4^ copies/L to 1.1E^+6^ copies/L corresponding to an input for library preparation ranging from 36 to 1,206 copies. A Spearman’s correlation showed a significant moderate positive correlation of mutation frequencies (r = 0.38, p = 0.002). The del60-70 mutation was not found by sequencing in eight samples despite including samples where Omicron BA.1 was the predominant variant and SARS-CoV-2 concentrations the highest, suggesting a lower sensitivity of this sequencing-based mutation monitoring. Results also showed important disparities in viral population dynamics over this period resulting from these datasets (Figure 6, panels B, C and D). The fluctuations observed in the mutation frequencies by sequencing have made curve smoothing difficult and the dynamics of change in majority populations were not evident to discern. Mutation frequencies for each sample and sequencing depth for each mutation position were available in supplementary data 3.

**Figure 6.**
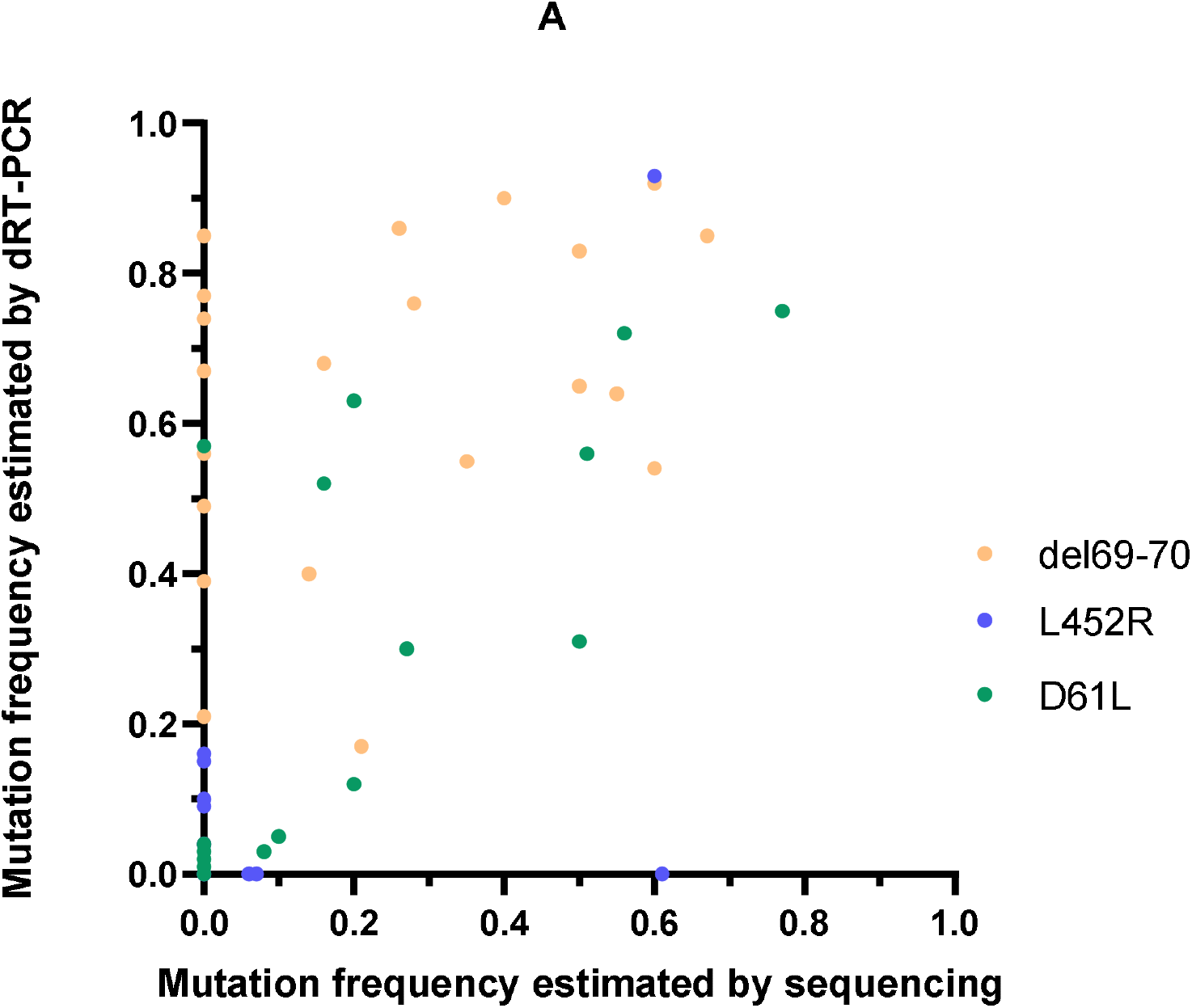
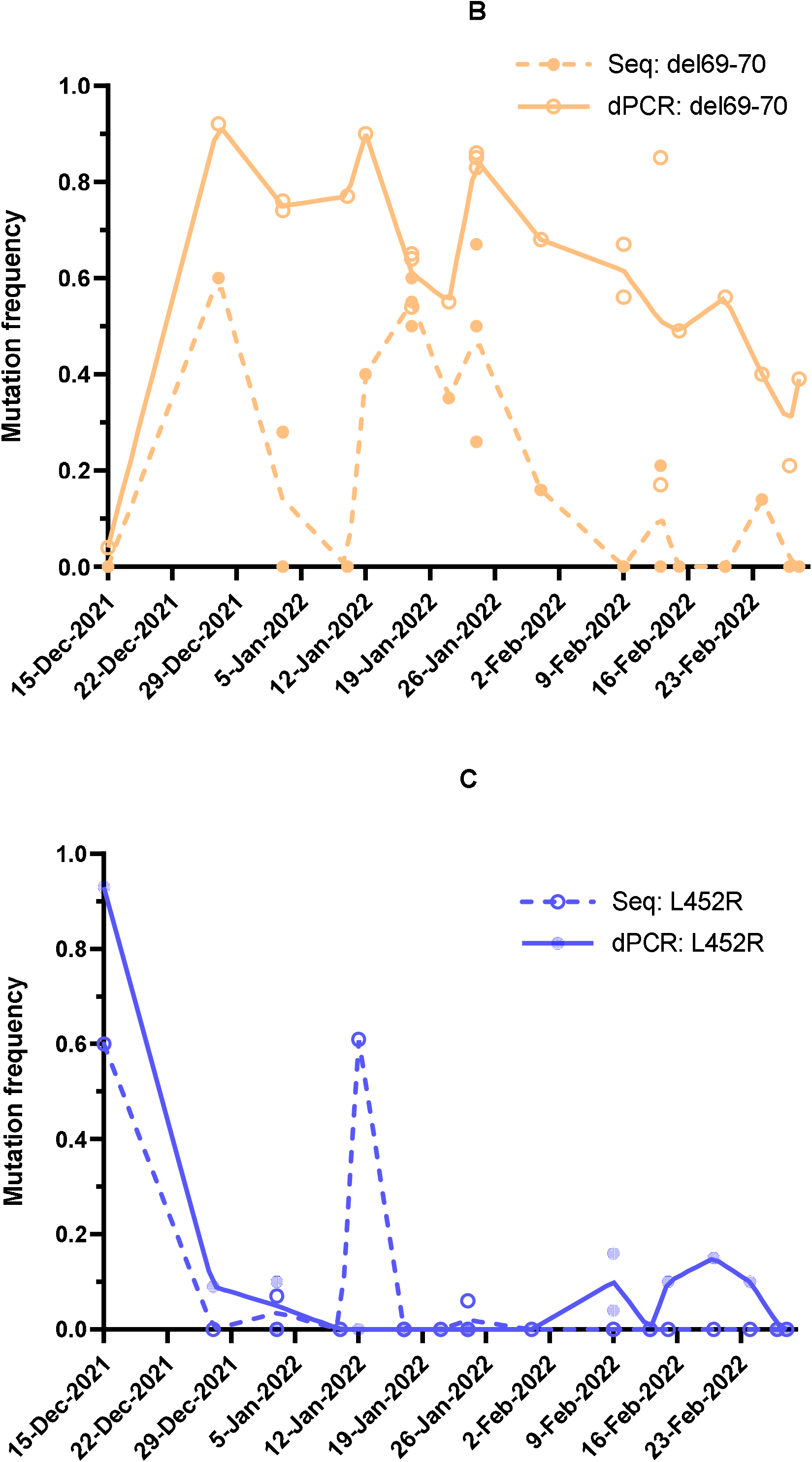
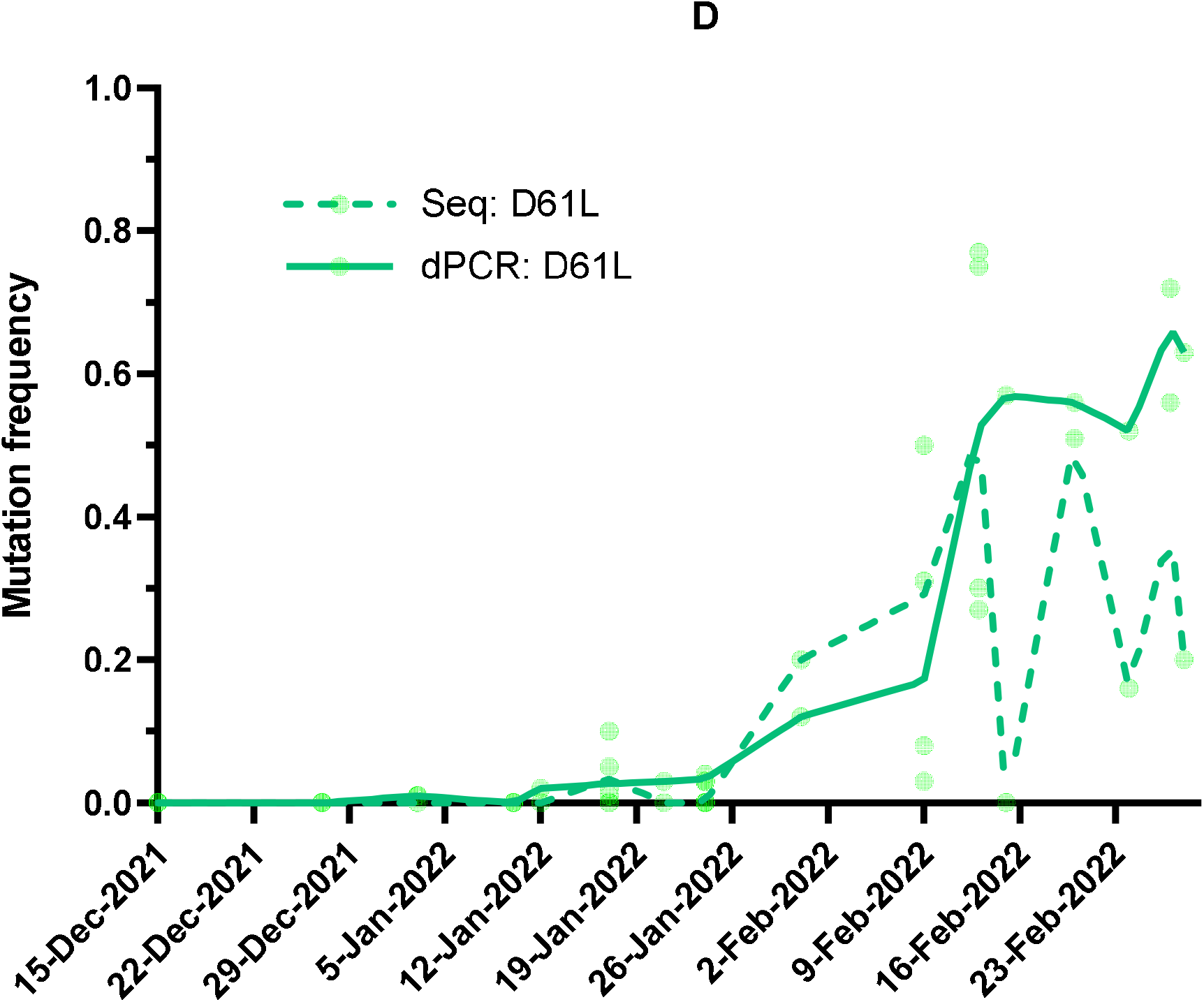
Comparison between mutation frequencies estimated by digital PCR and by sequencing in wastewater samples from december 15, 2021 to February 28, 2022. Panel A: Correlations estimated for the del69-70 (orange), L452R (blue) and D61L (green) Panel B: Evolutionary dynamics based on the del69-70 (orange) mutation frequency determined by digital PCR (solid line) or sequencing (dotted line) in wastewater. Panel C: Evolutionary dynamics based on the L452R (blue) mutation frequency determined by digital PCR (solid line) or sequencing (dotted line) in wastewater. Panel D: Evolutionary dynamics based on the D61L (green) mutation frequency determined by digital PCR (solid line) or sequencing (dotted line) in wastewater.

## Discussion

Omicron VOC is at the origin of an unprecedented upsurge in cases of COVID-19 since December 2021 in France. In Hong Kong, where people at risk (over 80 years old) are poorly vaccinated to date, a sharp increase in deaths has currently been observed (The New York Times, 2022). If Omicron VOC is spreading very rapidly in the population, the vaccination coverage in France (77,8% of the population with a complete vaccination schedule by March 15, 2022) may explain the lower severity of this VOC and its moderate impact on the French health system, compared to previous VOC. Viral spread in the general population may therefore become increasingly difficult to monitor with classical epidemiological indicators. Changes in tropism relative to Omicron VOC have also been reported. Nasopharyngeal swab-based screening may be less well suited to this new VOC or possible future variants, potentially leading to an underestimation of real viral circulation (Gupta, 2022; Marais et al., 2021).

Wastewater-based epidemiology allows tracking the overall dynamics of the SARS-CoV-2 epidemic (Medema et al., 2020b; Randazzo et al., 2020; Wurtzer et al., 2022, 2020). Variant identification results from sequencing, but such approach faces limitations for application in wastewater with respect to RNA quality and quantity (Lou et al., 2022), but also mathematical tools to deconvolute isolated mutations to reconstruct the original genomes in a mixture of variants (Agrawal et al., 2022b; Jahn et al., 2021; Smyth et al., 2022). Different studies have shown that it is also possible to monitor specific mutations suggestive of VOC to identify the emergence of VOC in a territory and to explain the predominance of certain VOC in epidemic waves (Caduff et al., 2022; Carcereny et al., 2022; Erster et al., 2022; Wurtzer et al., 2022). RT-qPCR approaches require calibration tools for absolute quantification or allelic discrimination (wild-type vs mutated) for relative quantification. Digital PCR allows more flexibility by offering a most probable number estimation (in accordance with Poisson’s law), by avoiding the realization of a standard curve and by allowing multiplexing of reactions.

The del69-70 mutation was shared by Alpha and Omicron BA.1 VOCs. However, Alpha VOC has been chronologically replaced by Delta VOC, to the point of disappearance as confirmed by the absence of del69-70 mutation detection since summer 2021 in wastewater. In this regard, the WHO no longer identifies Alpha VOC as a circulating VOC (World Health Organization, 2022). On the other hand, Omicron BA.1 VOC carries the synonymous mutation C21762T, the presence of which confers a better discrimination between the del69-70 mutation carried by Alpha VOC and that carried by Omicron BA.1 VOC. Detection of the del69-70 mutation resulting from Alpha VOC was therefore unlikely in winter 2021. To our knowledge, this study is the first to date the emergence of BA.1 and BA.2 sublineages of Omicron VOC in raw wastewater in France. The Omicron BA.1 VOC was detected earlier in the greater Paris area compared to other French regions. The Paris region is a transit hub between the different regions and the international. This observed precocity is therefore not surprising. The monitoring of specific mutations in wastewater has allowed the very early identification of the Omicron BA.1 VOC (Ferré et al., 2022) as well as the BA.2 sublineage from January 2022.

Retrospective studies have also demonstrated the presence of minor viral populations carrying the L452R mutation, suggestive of Delta VOC, as early as January 2021, in parallel with the strong expansion of Alpha VOC. The high transmissibility of Alpha VOC was at the origin of the third wave of COVID-19 encountered in France (Wurtzer et al., 2022). The introduction of vaccination coupled with acquired collective immunity to this VOC has probably allowed the expansion of an already circulating and better adapted variant such as Delta VOC whose L452R mutation allows a better escape to the immune response (Motozono et al., 2021). The results presented suggested that vaccination (and previous infections) could have contributed to the selection of the Delta VOC but also strongly reduced viral circulation with respect to the concentrations measured in the wastewater when the Delta VOC was in the majority. The large increase in viral genome concentration in wastewater during the Omicron BA.1 wave, compared with the Delta wave, was also consistent with the reduced efficacy of the Omicron VOC vaccine response (Cele et al., 2022). These results were consistent with a vaccine-induced reduction in severe forms of COVID-19 resulting from Omicron VOC infection without preventing viral circulation in the populations.

To date, VOCs are effectively monitored by conventional epidemiological indicators and intensive sequencing of strains isolated from patients. The dynamics of VOC-suggestive mutations in wastewater were consistent with VOC dynamics in patients, particularly with respect to the early detections, wave dynamics and changes in dominant populations. While variants may emerge simultaneously (e.g. Alpha and Delta VOCs), there were no periods in which different SARS-CoV-2 VOCs were codominant. Over the periods studied, the co-spreading of different variants in a population was finally only very transient and each variant was bound to be replaced by another one. This constant variant replacement demonstrated the SARS-CoV-2 evolution spreading within a population that has been largely immunized collectively either by one or more infections or by vaccination. Thus, maximum VOC replacement rates were calculated for variants that have circulated extensively in France since January 2021. Beta and Gamma VOCs represented at most 13.5% of the sequenced strains (April 26, 2021) and their detection in wastewater was not investigated in this study. The maximum VOC replacement rate for Alpha VOC (2,8%/day) was very close to the growth rate (4%/day) estimated in another study (Caduff et al., 2022). It was quite remarkable that the maximum replacement rate has progressively increased as new VOCs have emerged, suggesting increasingly contagious variants. Furthermore, the sum of the proportions of the different targeted mutations explained the majority of circulating SARS-CoV-2 genomes from November 2021 to March 2022. Contrary to the first studied period (from January to June 2021), the sum of the del69-70 and L452R mutations only partially explained the total circulating genomes. This result was consistent with the cumulative proportions of the other VOCs (e.g. Beta and Gamma VOCs) and the decreasing proportion of the original strain of SARS-CoV-2, and could underline the increasing “fitness” of circulating VOC.

In addition to the method variability (Bivins et al., 2021b, 2021a) and potentially different fecal shedding depending on the variants, the SARS-CoV-2 concentrations in wastewater can be affected by heavy rainfall and population movement. Mathematical algorithms have been proposed to compare data sets obtained from different methods or laboratories, and to correct the raw concentrations by the flow of wastewater inlet (Cluzel et al., 2022; Courbariaux et al., 2022). The greater Paris area is particularly affected by the latter because it is a very touristy region, and this region concentrates a very strong professional activity. This second period was strongly impacted by the school vacations of Christmas (from December 20, 2021 to January 1, 2022) and winter (from February 19 to March 7, 2022) during which many people leave the region. To account for these population variations, PMMoV genome, a human biomarker, was also measured by digital RT-PCR as well as the daily drinking water consumption in Paris. The PMMoV is a plant virus that is very abundant in human stools, although the origin of this presence is not really known. Identified as plant virus (pepper), food is suspected to be the main source of contamination (Colson et al., 2010). Its detection has been proposed as tracer of anthropogenic activity (D’Aoust et al., 2021; Malla et al., 2019). In this study, the PMMoV genome concentration in raw wastewater showed an important variability that did not highlight the population movements in the studied watershed as demonstrated by drinking water consumption and vacation calendar. Using population indicator such as drinking water consumption or PMMoV shedding, the normalized viral dynamics in wastewater were quite different. Normalization using drinking water consumption generated a curve very close to the dynamics based on raw concentrations, with a more precise definition of the peak of the curve. The correlation of these results with the regional incidence curve was also the best fitted. The overall dynamic revealed a temporal advance in agreement with other results (Caduff et al., 2022; Cluzel et al., 2022; Wurtzer et al., 2022). Nevertheless, this temporal advance on the wave related to the Omicron VOC appeared to be greater than the advance reported for previous VOCs. Similar advance can be observed on SARS-CoV-2 wastewater monitoring data in the U.S (Biobot.io, 2022). This finding could be explained by a higher proportion of asymptomatic or paucisymptomatic infections that would be detected later. Normalization based on PMMoV genome concentration suggested a wave of viral genomes in wastewater synchronized with incidence. Surprisingly, this approach showed a viral rebound in February 2022 that was not observed on the other modeled curves, nor on the incidence curve. This observation was probably the reason for the lower correlation. If other human biomarkers could be evaluated such as CrAssphages or specific F-RNA phages, a previous study also concluded that normalization of SARS-CoV-2 signal by fecal indicator did not improve the correlation with the incidence (Ai et al., 2021). All three approaches agreed on a significant resurgence of the viral circulation observed in wastewater in early March 2022 confirmed by incidence data. The circulation of variants leading to less severe infections, resulting either from acquired collective immunity (vaccination and infections) or from an attenuation of the intrinsic viral pathogenicity, could lead to a less good identification of the cases of infection in the populations, and thus to an apparent incidence decorrelated from the real incidence. In this sense, viral monitoring in wastewater will allow an unbiased approach to viral circulation.

This study also demonstrated the interest of monitoring specific mutations evocative of VOC because of the simplicity of setting up such an measurement, its low cost and the result quickness, which are indispensable for informing health authorities. This study also highlighted the limitations of sequencing in wastewater with respect to analytical sensitivity that may result from the low concentration of SARS-CoV-2 genomes, the very low concentration of mutations in minority variant populations, insufficient sequencing depth, and non-homogeneous coverage along the entire viral genome. Similar limitations have been already reported (Caduff et al., 2022). The accumulation of mutations identified by sequencing is also challenged by the reconstruction of the genomes from which these mutations originate. Mathematical algorithms for deconvolution of isolated mutations or long fragment sequencing approaches to identify different polymorphisms in the same reads may improve the predictivity of sequencing. To date, sequencing must be interpreted with caution for quantitative assessments of mutation frequency in wastewater.

## Conclusion

The quantification of mutations suggestive of VOCs circulating in wastewater has allowed a depiction of the VOC propagation dynamics in the greater Paris area population. This approach also allowed early detection of the Omicron VOC BA.1 and BA.2 lineages emergence in this area. The observed dynamics confirmed that the vaccination strategy of the human population, as well as exposition of previous variants, probably contributed to the selection of Delta VOC to the detriment of Alpha VOC, but also to the strong decrease of the viral circulation in the human population during the same period, confirming the effectiveness of the vaccine response against these VOCs. Moreover, the normalization of concentrations based on the drinking water consumption in Paris seemed to be a relevant method to follow the population movements in this region. This normalized approach provided a viral dynamic in close relationship with the regional incidence curve. Spreading of variants leading to less severe infections could also lead to a detection delay through patient-centered syndromic surveillance tools. Thus, raw wastewater testing will still allow this tracking, as well as the VOC dynamics monitoring during the spread of SARS-CoV-2 in populations.

## Data Availability

All data produced in the present study are available upon reasonable request to the authors

## Acknowledgments

We thank all the people who have made it possible to collect all these wastewater samples since the beginning of the pandemic, especially the team of sewage workers of Paris, France, who set up a specific monitoring. Technical teams of WWTP (SIAM, SIAAP and other utilities) should also be thanked. We also thank all the people who participated in the exchange of ideas and who cannot be listed as co-authors of the study.

^*^ member of the scientific interest group OBEPINE : IRBA (Boni M), Eau de Paris (Wurtzer S, Moulin L), Sorbonne Université (Mouchel JM, Maday Y, Marechal V), IFREMER (Le Guyader S), LCPME (Bertrand I, Gantzer C).

---

**Supplementary data 1.**
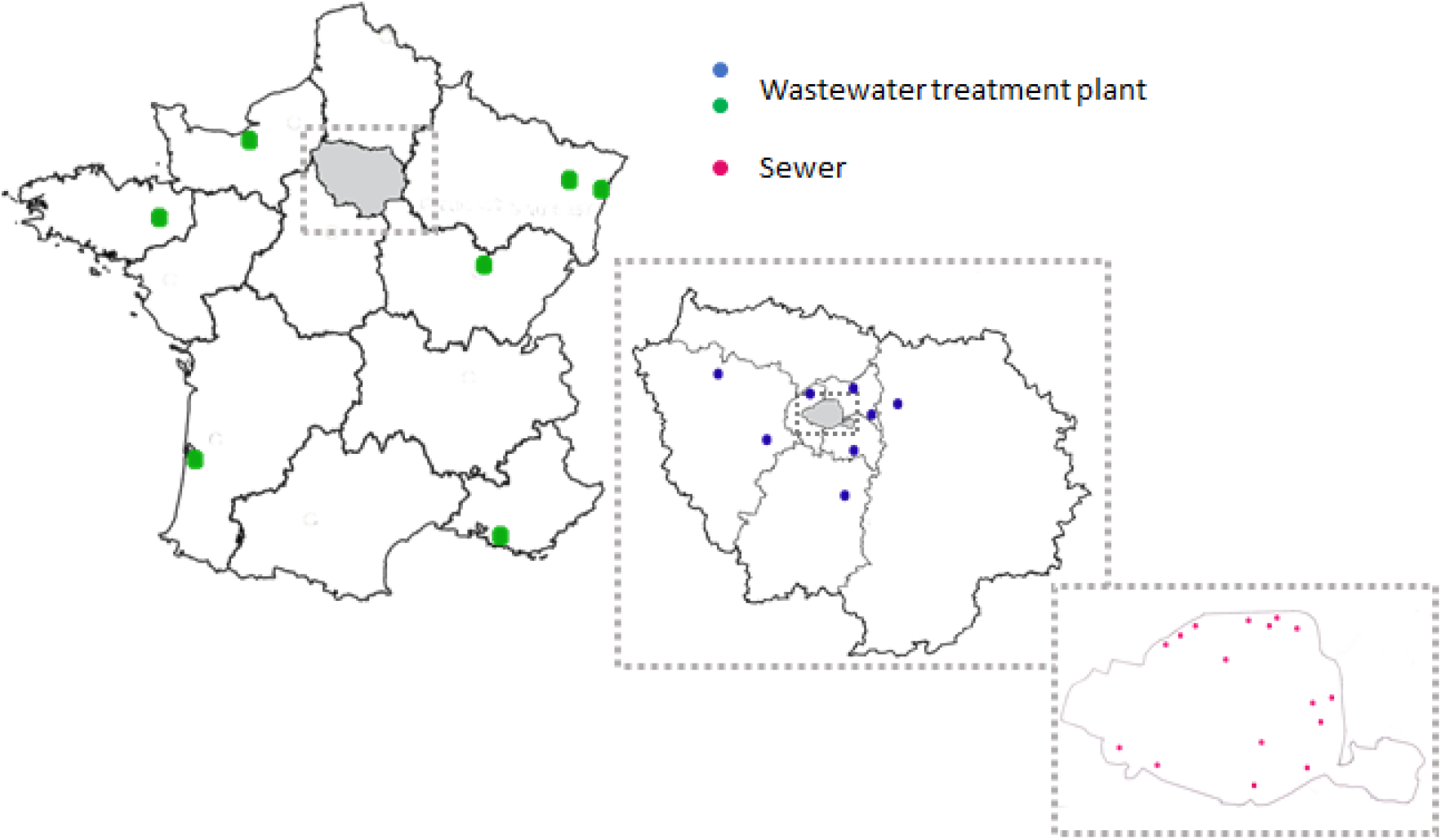
Location of wastewater treatment plants or sewer sampled.

**Supplementary data 2.**
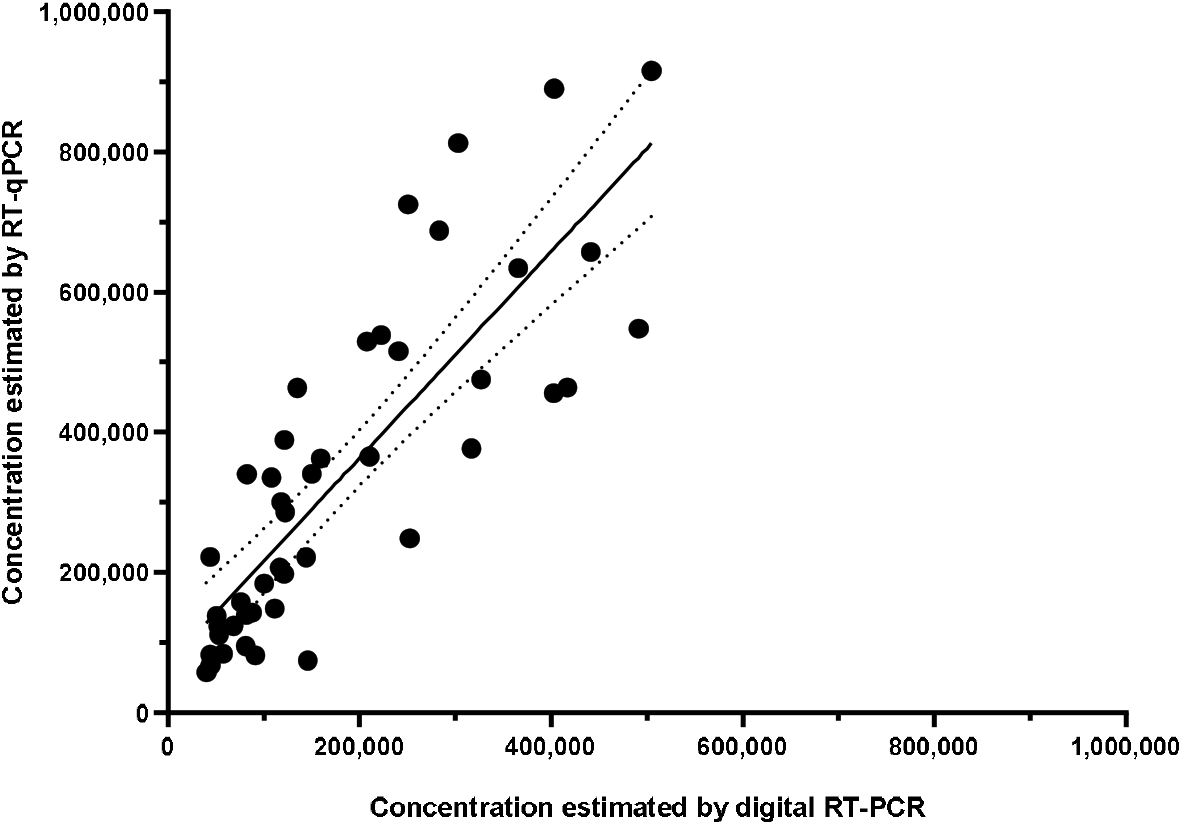
Comparison between quantification by digital RT-PCR and RT-qPCR

**Supplementary data 3.**
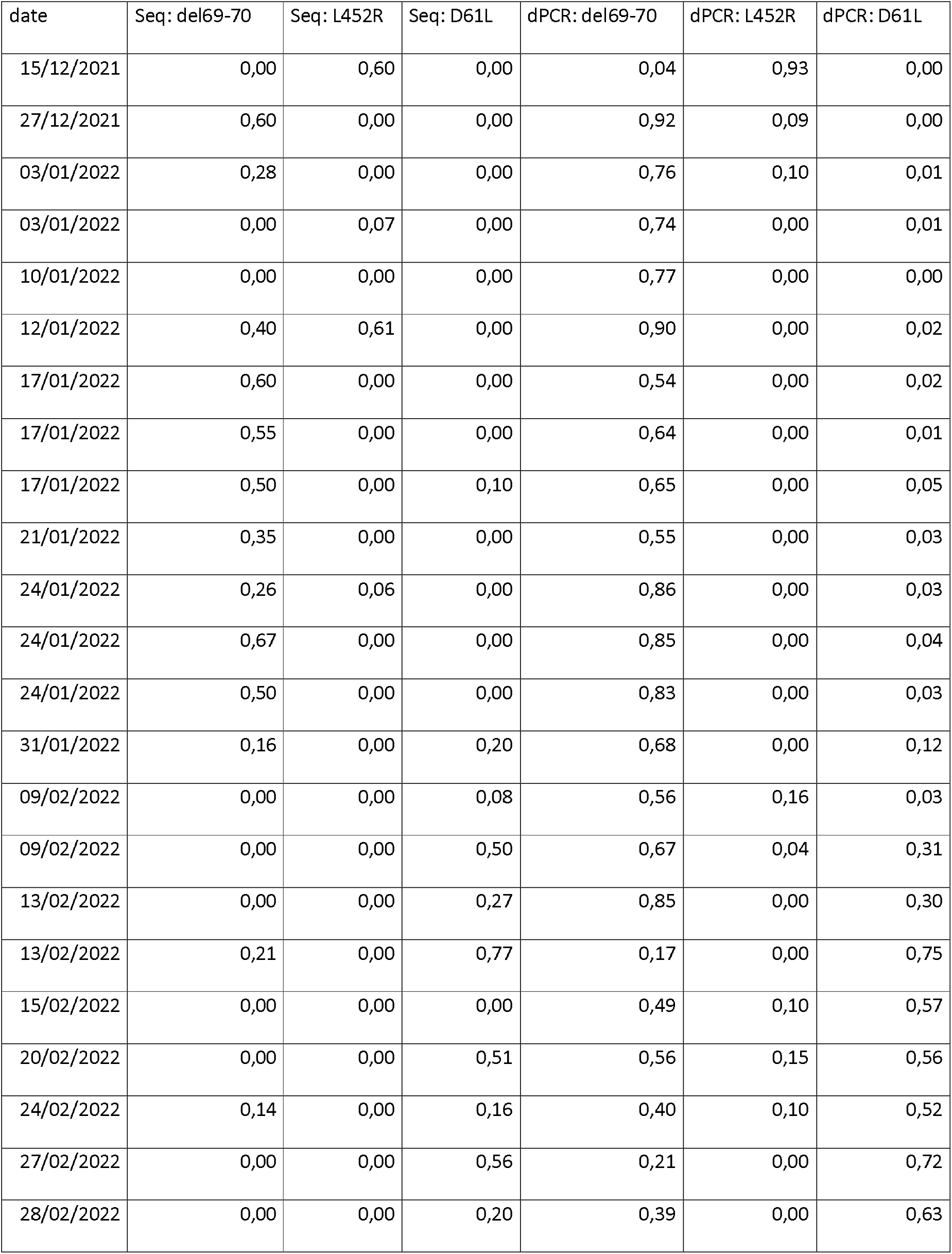
Mutation frequencies estimated by sequencing or digital RT-PCR (per sample) Depth sequencing for specific mutation locations.

| date | Seq: del69-70 | Seq: L452R | Seq: D61L | dPCR: del69-70 | dPCR: L452R | dPCR: D61L |
| --- | --- | --- | --- | --- | --- | --- |
| 15/12/2021 | 0,00 | 0,60 | 0,00 | 0,04 | 0,93 | 0,00 |
| 27/12/2021 | 0,60 | 0,00 | 0,00 | 0,92 | 0,09 | 0,00 |
| 03/01/2022 | 0,28 | 0,00 | 0,00 | 0,76 | 0,10 | 0,01 |
| 03/01/2022 | 0,00 | 0,07 | 0,00 | 0,74 | 0,00 | 0,01 |
| 10/01/2022 | 0,00 | 0,00 | 0,00 | 0,77 | 0,00 | 0,00 |
| 12/01/2022 | 0,40 | 0,61 | 0,00 | 0,90 | 0,00 | 0,02 |
| 17/01/2022 | 0,60 | 0,00 | 0,00 | 0,54 | 0,00 | 0,02 |
| 17/01/2022 | 0,55 | 0,00 | 0,00 | 0,64 | 0,00 | 0,01 |
| 17/01/2022 | 0,50 | 0,00 | 0,10 | 0,65 | 0,00 | 0,05 |
| 21/01/2022 | 0,35 | 0,00 | 0,00 | 0,55 | 0,00 | 0,03 |
| 24/01/2022 | 0,26 | 0,06 | 0,00 | 0,86 | 0,00 | 0,03 |
| 24/01/2022 | 0,67 | 0,00 | 0,00 | 0,85 | 0,00 | 0,04 |
| 24/01/2022 | 0,50 | 0,00 | 0,00 | 0,83 | 0,00 | 0,03 |
| 31/01/2022 | 0,16 | 0,00 | 0,20 | 0,68 | 0,00 | 0,12 |
| 09/02/2022 | 0,00 | 0,00 | 0,08 | 0,56 | 0,16 | 0,03 |
| 09/02/2022 | 0,00 | 0,00 | 0,50 | 0,67 | 0,04 | 0,31 |
| 13/02/2022 | 0,00 | 0,00 | 0,27 | 0,85 | 0,00 | 0,30 |
| 13/02/2022 | 0,21 | 0,00 | 0,77 | 0,17 | 0,00 | 0,75 |
| 15/02/2022 | 0,00 | 0,00 | 0,00 | 0,49 | 0,10 | 0,57 |
| 20/02/2022 | 0,00 | 0,00 | 0,51 | 0,56 | 0,15 | 0,56 |
| 24/02/2022 | 0,14 | 0,00 | 0,16 | 0,40 | 0,10 | 0,52 |
| 27/02/2022 | 0,00 | 0,00 | 0,56 | 0,21 | 0,00 | 0,72 |
| 28/02/2022 | 0,00 | 0,00 | 0,20 | 0,39 | 0,00 | 0,63 |

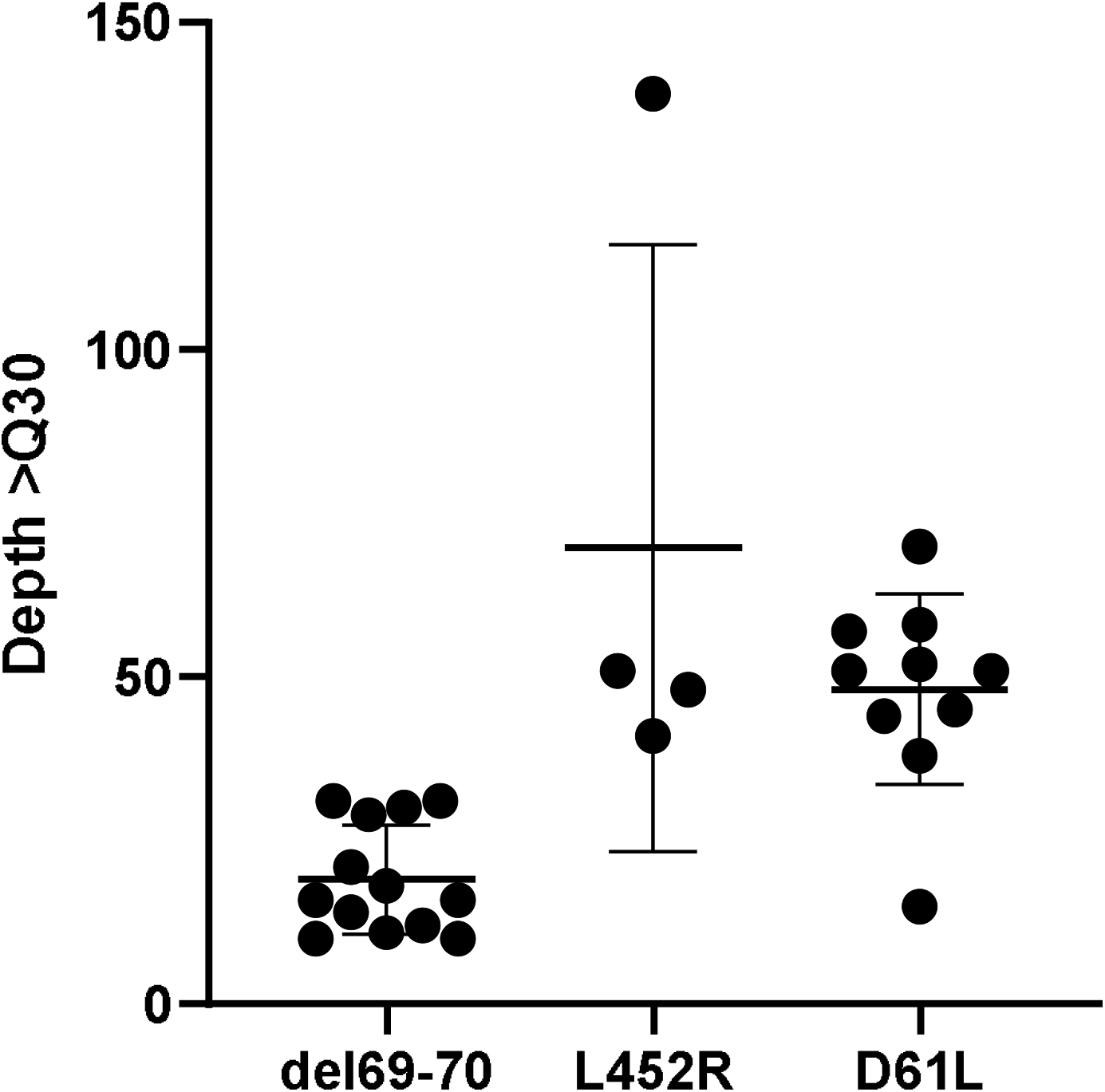

